# Genome-wide association study analysis of disease severity in Acne reveals novel biological insights

**DOI:** 10.1101/2023.11.13.23298473

**Authors:** Zhaohui Du, Tejaswi Iyyanki, Samuel Lessard, Michael Chao, FinnGen, Christian Asbrand, Dany Nassar, Katherine Klinger, Emanuele de Rinaldis, Shameer Khader, Clément Chatelain

## Abstract

Acne vulgaris is a common skin disease that affects >85% of teenage young adults among which >8% develop severe lesions that leaves permanent scars. Genetic heritability studies of acne in twin cohorts have estimated that the heritability for acne is 80%. Previous genome-wide association studies (GWAS) have identified 50 genetic *loci* associated with increased risk of developing acne when compared to healthy individuals. However only a few studies have investigated genetic association with disease severity. GWAS of disease progression may provide a more effective approach to unveil potential disease modifying therapeutic targets.

Here, we performed a multi-ethnic GWAS analysis to capture disease severity in acne patients by using individuals with normal acne as a control. Our cohort consists of a total of 2,956 participants, including 290 severe acne cases and 930 normal acne controls from FinnGen, and 522 cases and 1,214 controls from BioVU. We also performed mendelian randomization (MR), colocalization analyses and transcriptome-wide association study (TWAS) to identify putative causal genes. Lastly, we performed gene-set enrichment analysis using MAGMA to implicate biological pathways that drive disease severity in Acne.

We identified two new loci associated with acne severity at the genome-wide significance level, six novel associated genes by MR, colocalization and TWAS analyses, including genes *CDC7, SLC7A1, ADAM23, TTLL10, CDK20* and *DNAJA4*, and 5 novel pathways by MAGMA analyses. Our study suggests that the etiologies of acne susceptibility and severity have limited overlap, with only 26% of known acne risk loci presenting nominal association with acne severity and none of the novel severity associated genes reported as associated with acne risk in previous GWAS.

## Introduction

Acne vulgaris is a chronic inflammatory dermatosis that is characterized by comedones and inflammatory lesions including papules, pustules, and nodules^1^. It is the most common skin disorder that affects >85% of teenage young adults where >8% develop severe lesions that leaves permanent scars^2^. Severe acne may have a significant long-term negative impact on emotional and mental health including low self-esteem and social isolation^3^. The pathogenesis of acne involves a complex interplay between sebum production, follicular keratinisation, inflammation, and colonization of pilosebaceous follicles by Propionibacterium acnes^1^.

Genetic heritability studies of acne in twin cohorts have shown that the heritability for acne is 80%^4^. Previous genome-wide association studies (GWAS) have identified 50 genetic *loci* associated with increased risk of developing acne when compared to healthy individuals among Caucasian or Asian populations^5–9^. However only a few studies have investigated genetic associations with disease severity. Whether biological mechanisms driving disease onset and progression are the same is still an open question in human genetics, as for many complex diseases. GWAS that focus on disease severity or progression may provide a more effective approach to unveil potential disease modifying therapeutic targets. Such studies have recently become possible with the rise of large-scale biobanks, such as FinnGen and UK Biobank, combining genetic and electronic health records.

Here, we performed a multi-ethnic GWAS analysis to capture disease severity in acne patients by using individuals with normal acne as a control. Our cohort consists of a total of 2,956 participants, including 290 severe acne cases and 930 normal acne controls from FinnGen release 10, and 522 cases and 1,214 controls from the BioVU biorepository extracted by Nashville Biosciences. We also performed mendelian randomization (MR), colocalization analyses and transcriptome-wide association study (TWAS) to identify putative causal genes. Lastly, we performed gene-set enrichment analysis using MAGMA to implicate biological pathways that drive disease severity in acne.

## Results

### GWAS analyses

We discovered one novel locus at 7q22.3 significantly associated with acne severity at the genome-wide significance level (P-value < 5 × 10^−8^) among participants with African ancestry (AA) in the BioVU cohort (Figure 1, Supplementary figure 1). The lead variant, rs115325598 (OR = 8.22; 95% CI: 3.72, 18.13; P-value = 3.15 × 10^−8^), is located at the intron region of gene *ATXN7L1*. The effect allele (C) of rs115325598 has a frequency of 6% in controls and 16% in severe acne cases among AA population in BioVU. There is one additional genome-wide significant locus at 12p12.2 among participants with European ancestry (EA) in BioVU cohort (supplementary figure 2). The lead variant, rs116779562 (OR = 11.04; 95% CI: 4.61, 26.39; P-value = 3.25 × 10^−8^), located at an intergenic region between genes *LINC02398* and *PDE3A*. However, this association was not replicated among FinnGen population (OR = 1.49; 95% CI: 0.47, 4.77; P-value = 0.50). The effect allele (T) of this variant is rare in controls (0.3%) and has low frequency among severe acne cases (2%). No genome-wide signal was detected in any other GWAS analyses (Supplementary figure 1).

**Figure 1:**
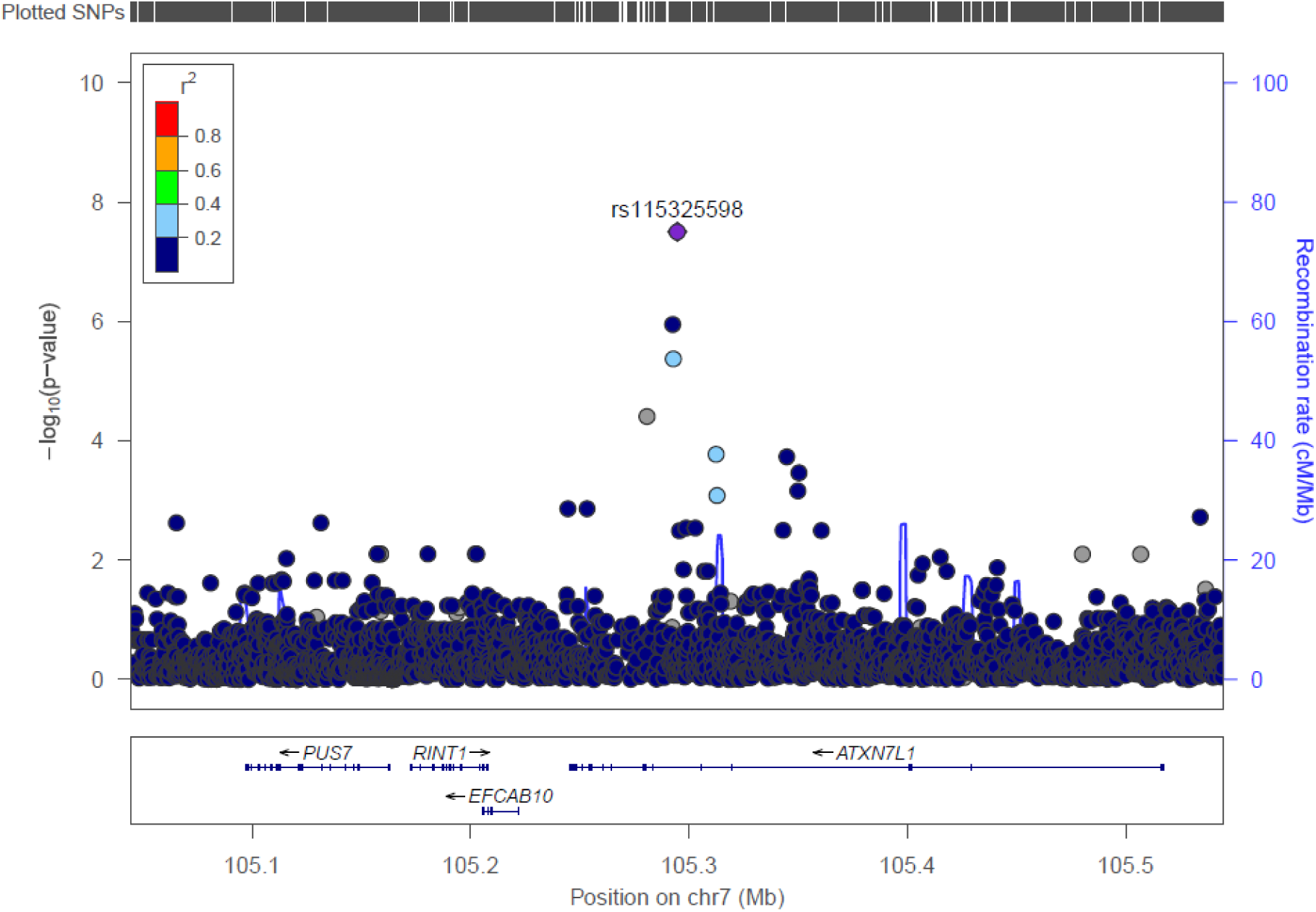
Regional association plots of the 7q22.3 risk region for acne severity among African ancestry (AA) population in BioVU cohort. Single-nucleotide polymorphisms (SNPs) are plotted by position (x-axis) and −log10 p-value (y-axis). r^2^ was estimated from AA individuals in phase III 1000 Genomes Project (1KGP) data. The most statistically significant associated SNP (purple diamond) at 7q22.3 is rs115325598. The surrounding SNPs are colored to indicate pairwise correlation with the index SNP.

### Variant annotation

We annotated the impact of all suggestive variants with *P*<1x10^-6^ including coding variants and predicted gain or loss of function variants (GoLoF), and also mapped these variants to genes using activity-by-contact (ABC) enhancer-promoter interactions^10^. Among the 5 single and meta-analyses GWASs, a total of 38 unique variants passed suggestive significance level. Two variants, rs3742185 and rs186795319 are annotated as coding variants of genes *ANKRD10* and *UMODL1*, with predicted functional consequences being missense and stop gained, respectively (Supplementary table 1).

### Mendelian randomization and Colocalization analyses

In MR analyses, 36 genes were identified as significantly associated with acne severity at q-value < 0.05. We then conducted a colocalization analysis to further assess the probability that severe acne associated SNPs and eQTLs share the same causal genetic variant. Six genes (*ADAM23, CDC7, CDK20, DNAJA4, SLC7A1, TTLL10*) were found to have a colocalization probability ≥ 0.8 in at least one tissue/cell type. (Table 1, Supplementary table 2).

**Table 1.** Genes that are associated with severe acne risk in the MR (Q-value < 0.05) and Colocalization analyses (PP.H4 ≥ 0.8).

| Gene | GWAS | Region | MR beta* | MR q-value* | No. tissues/ cell types with Coloc PP.H4 $\geq 0.8$ | Max. Coloc PP.H4* | Tissue/cell type with Max. Coloc PP.H4* |
| --- | --- | --- | --- | --- | --- | --- | --- |
| *ADAM23* | BioVU EA | chr2: 207062772-207562772 | 1.29 | $5.92 \times 10^{-3}$ | 2 | 0.97 | Sigmoid colon |
| *CDC7* | BioVU EA | chr1: 91716406-92216406 | 1.29 | $1.26 \times 10^{-3}$ | 13 | 1.00 | Monocyte |
| *CDK20* | BioVU EA | chr9: 90214701-90714701 | -8.15 | $1.61 \times 10^{-3}$ | 1 | 0.93 | Brain |
| *DNAJA4* | BioVU EA + FinnGen | chr15: 78315188-78815188 | 0.32 | $9.86 \times 10^{-3}$ | 2 | 1.00 | Monocyte |
| *SLC7A1* | BioVU EA | chr13: 30017359-30517359 | 4.88 | $8.06 \times 10^{-4}$ | 4 | 0.97 | Blood |
| *SLC7A1* | BioVU EA + FinnGen | chr13: 30018119-30518119 | 3.50 | $8.89 \times 10^{-3}$ | 3 | 0.96 | Blood |
| *SLC7A1* | BioVU EA + AA + FinnGen | chr13: 30017359-30517359 | 3.05 | 0.02 | 1 | 0.97 | Blood |
| *TTLL10* | BioVU EA | chr1: 816097-1316097 | -1.34 | 0.04 | 1 | 0.87 | Stomach |

\*Displayed results in tissues with MR Q-value $< 0.05$ and with the highest colocalization PP.H4.

### Gene-set analyses using MAGMA identified pathways associated with severe acne risk

In analyses using MAGMA, no associated gene was identified in the gene level analyses for severe acne risk. Subsequent gene-set analysis demonstrated significant enrichment in four pathways/gene sets at FDR < 0.05 (Table 2). The top pathway is “response to fluoride”, which includes five genes, including *COL1A1, PLCB1, ATG5, PON1* and *ASPN*, that collectively regulate the change in state or activity of a cell or an organism (in terms of movement, secretion, enzyme production, gene expression, etc.) as a result of a fluoride stimulus.

**Table 2.** Pathways statistically significantly associated with acne severity in MAGMA analysis.

| Pathway Name | Discovery GWAS | P-value | Number of genes |
| --- | --- | --- | --- |
| GOBP: response to fluoride | BioVU EA + FinnGen | $1.72 \times 10^{-7}$ | 5 |
| GOCC: ISWI type complex | BioVU EA + FinnGen | $1.12 \times 10^{-6}$ | 11 |
| GOCC: intercellular bridge | BioVU EA + AA + FinnGen | $1.59 \times 10^{-6}$ | 67 |
| GOBP: sperm DNA condensation | BioVU EA | $1.23 \times 10^{-6}$ | 10 |

### Potential pathogenic gene prioritization

After getting the list of candidate pathogenic genes associated with severe acne risk using GWAS (the nearest gene of the genome-wide significant risk variant), MR and colocalization analyses, we further gathered additional evidences that may support the pathogenicity of those genes through evaluating genetic associations with other diseases in a phenome-wide search using an in-house gene prioritization scheme, differential expressions in other diseases across multiple tissues/cell types, bulk and single cell gene expression levels by tissues and cell types. The credentialing results for each gene are summarized below (Table 3, Supplementary table 3-5, Supplementary figures 3-4).

**Table 3:**
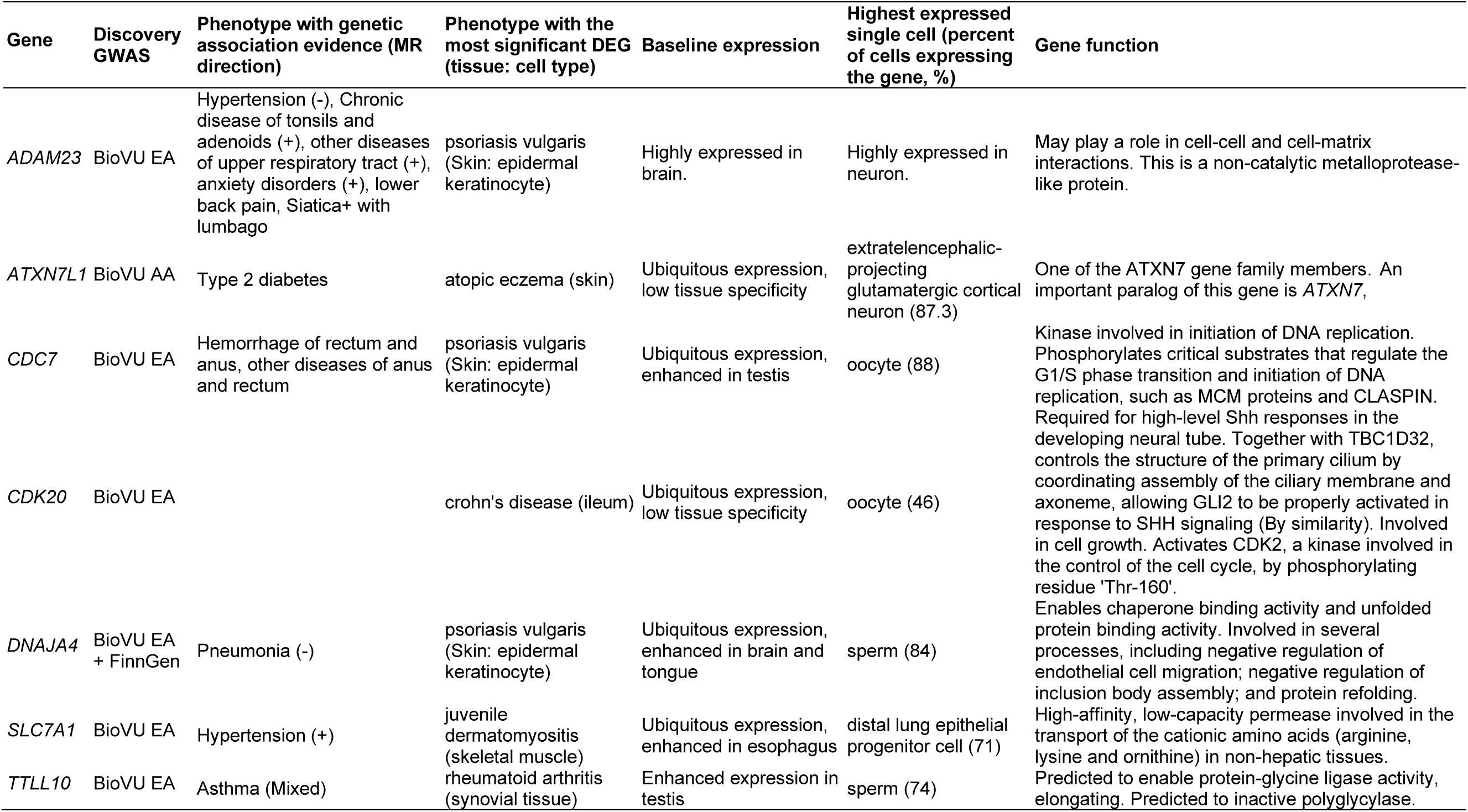
Summary of genetic-/transcriptomic-associated phenotypes and the highest expression tissues/cell types of candidate pathogenic genes.

### *ADAM23* (ADAM Metallopeptidase Domain 23)

In genetic prioritization ranking with other potential diseases, *ADAM23* showed evidence of association with a variety of diseases including hypertension, upper respiratory tract diseases (Supplementary table 3). In differential gene expression analysis, *ADAM23* is shown to be mostly upregulated in epidermal keratinocyte cell in skin tissue of psoriasis vulgaris (log2-fold change = 2.8, adjusted P=9.4x10^-18^) and downregulated in CD14+ monocyte cell in skin tissue of clinically isolated syndrome (log2-fold change = -3.7, adjusted P=1.4x10^-13^) (Supplementary figure 3, Supplementary table 4). In baseline gene expression data, *ADAM23* is mostly expressed in brain and heart, followed by CD4-positive, alpha-beta thymocyte cells (Supplementary figure 4). In single cell data, *ADAM23* is most expressed in neural cells, with 99.1% of extratelencephalic-projecting glutamatergic cortical neuron cells expressing the gene, followed by corticothalamic-projecting glutamatergic cortical neuron (97.7%) (Supplementary table 5).

### *ATXN7L1* (Ataxin 7 Like 1)

In genetic prioritization ranking with other potential related diseases, *ATXN7L1* showed evidence of association with type 2 diabetes, with the instrument variant being a lead coding variant in this gene (Supplementary table 3). In differential gene expression analysis, *ATXN7L1* is shown to be mostly upregulated in neutrophil cell in blood of septic arthritis (log2-fold change = 1.3, adjusted P=6.6x10^-8^) and downregulated in epidermal keratinocyte cell in skin tissue of psoriasis vulgaris (log2-fold change = -1.6, adjusted P=7.3x10^-15^) (Supplementary figure 3, Supplementary table 4). *ATXN7L1* is ubiquitously expressed across multiple tissues with low tissue specificity (Supplementary figure 4). In single cell data, *ATXN7L1* is most expressed in extratelencephalic-projecting glutamatergic cortical neuron, with 87.3% of cells expressing this gene (Supplementary table 5).

### *CDC7* (Cell Division Cycle 7)

In genetic prioritization ranking with other potential related diseases, *CDC7* showed evidence of association with hemorrhage of rectum and anus (Supplementary table 3). In differential gene expression analysis, *CDC7* is shown to be mostly upregulated in epidermal keratinocyte cell of skin tissue in psoriasis vulgaris (log2-fold change = 4.2, adjusted P=6.7x10^-18^) and downregulated in platelet of colorectal cancer (log2-fold change = -2.5, adjusted P=1.0x10^-4^) (Supplementary figure 3, Supplementary table 4). *CDC7* is broadly expressed in testis, lymph node, bone marrows and multiple other tissues (Supplementary figure 4). In single cell data, *CDC7* is most expressed in germ cell, with 87.9% of oocyte cells expressing this gene Supplementary table 5).

### *CDK20* (Cyclin Dependent Kinase 20)

*CDK20* did not show genetic evidence of association with any disease using the genetic prioritization ranking scheme. In differential gene expression analysis, *CDK20* is shown to be mostly upregulated in skin tissue of atopic dermatitis (log2-fold change = 1.1, adjusted P=4.0x10^-2^) and downregulated in PBMC of clinically isolated syndrome (log2-fold change = -1.5, adjusted P=1.0x10^-12^) (Supplementary figure 3, Supplementary table 4). *CDK20* is ubiquitously expressed across multiple tissues with low tissue specificity (Supplementary figure 4). In single cell data, *CDK20* is most expressed in germ cell, with 45.6% of oocyte cells expressing this gene (Supplementary table 5).

### *DNAJA4* (DnaJ Heat Shock Protein Family (Hsp40) Member A4)

In genetic prioritization ranking with other potential related diseases, *DNAJA4* showed evidence of association with pneumonia, with a significant negative MR (MR beta = - 0.03) association with plasma pQTL (Supplementary table 3). In differential gene expression analysis, *DNAJA4* is shown to be mostly upregulated in cytotoxic T cell in blood of asthma (log2-fold change = 1.7, adjusted P=4.7x10^-5^) and downregulated in epidermal keratinocyte cell in skin tissue of psoriasis vulgaris (log2-fold change = -3.7, adjusted P=2.4x10^-18^) (Supplementary figure 3, Supplementary table 4). *DNAJA4* is broadly expressed in bone marrow, brain, heart, skeletal muscle tissue and other tissues (Supplementary figure 4). In single cell data, *DNAJA4* is most expressed in germ cell, with 84.5% of sperm cells expressing this gene (Supplementary table 5).

### *SLC7A1* (Solute Carrier Family 7 Member 1)

In genetic prioritization ranking with other potential related diseases, *CDC7* showed evidence of association with hypertension, with a significant positive MR (MR beta = 0.11∼0.28, q-value < 0.05) association and high colocalization (maximum PP.H4 = 0.99) in adrenal gland (Supplementary table 3). In differential gene expression analysis, *SLC7A1* is shown to be mostly upregulated in epidermal keratinocyte cell in skin tissue of psoriasis vulgaris (log2-fold change = 1.6, adjusted P=2.1x10^-17^) and downregulated in the PBMC in relapsing-remitting MS (log2-fold change = -1.9, adjusted P=9.9x10^-21^) (Supplementary figure 3, Supplementary table 4). *SLC7A1* is ubiquitously expressed in mucosa tissue in, transformed skin fibroblast, EBV-Transformed lymphocyte and other tissues (Supplementary figure 4). In single cell data, *SLC7A1* is most expressed in distal lung epithelial progenitor cell, with 71.2% of cells expressing this gene, followed by syncytiotrophoblast cell (65.8%) (Supplementary table 5).

### *TTLL10* (Tubulin Tyrosine Ligase Like 10)

In genetic prioritization ranking with other potential related diseases, *TTLL10* showed evidence of association with asthma, with the instrument variant of MR being a lead coding variant of this gene, a significant positive MR association and high colocalization (MR beta = 0.07, maximum PP.H4 = 0.99) in Brain Putamen basal ganglia cell, and a significant negative MR association and high colocalization in iPSC (MR beta = -0.07, maximum PP.H4 = 0.98, Supplementary table 3). It also has moderate evidence of association with hypothyroidism, with positive MR association (MR beta = 0.38) and high colocalization (PP.H4 = 0.89) in whole blood. In differential gene expression analysis, *TTLL10* is shown to be mostly upregulated in skin tissue of psoriasis (log2-fold change = 1.3, adjusted P=4.4x10^-2^) and downregulated in plasma blast cell of systemic lupus erythematosus (log2-fold change = -1.8, adjusted P=3.7x10^-3^) (Supplementary figure 3, Supplementary table 4). In baseline gene expression, *TTLL10* is mostly expressed in the testis tissue (Supplementary figure 4). In single cell data, *TTLL10* is most expressed in sperm, with 74.5% of cells expressing this gene (Supplementary table 5).

### Evaluation of known acne risk *loci* in severe versus normal acne comparisons

We extracted a total of 66 unique SNP associations from all 5 GWASs for the 50 reported acne risk *loci*. Among the 66 SNPs, 17 (26%) were nominally significant (P-value < 0.05) in any of the current GWASs, with the lowest P-value being 3.6x10^-3^ at rs17692425 in FinnGen GWAS; however, no SNP remained significant after Bonferroni correction (Supplementary table 6, figure 2).

**Figure 2.**
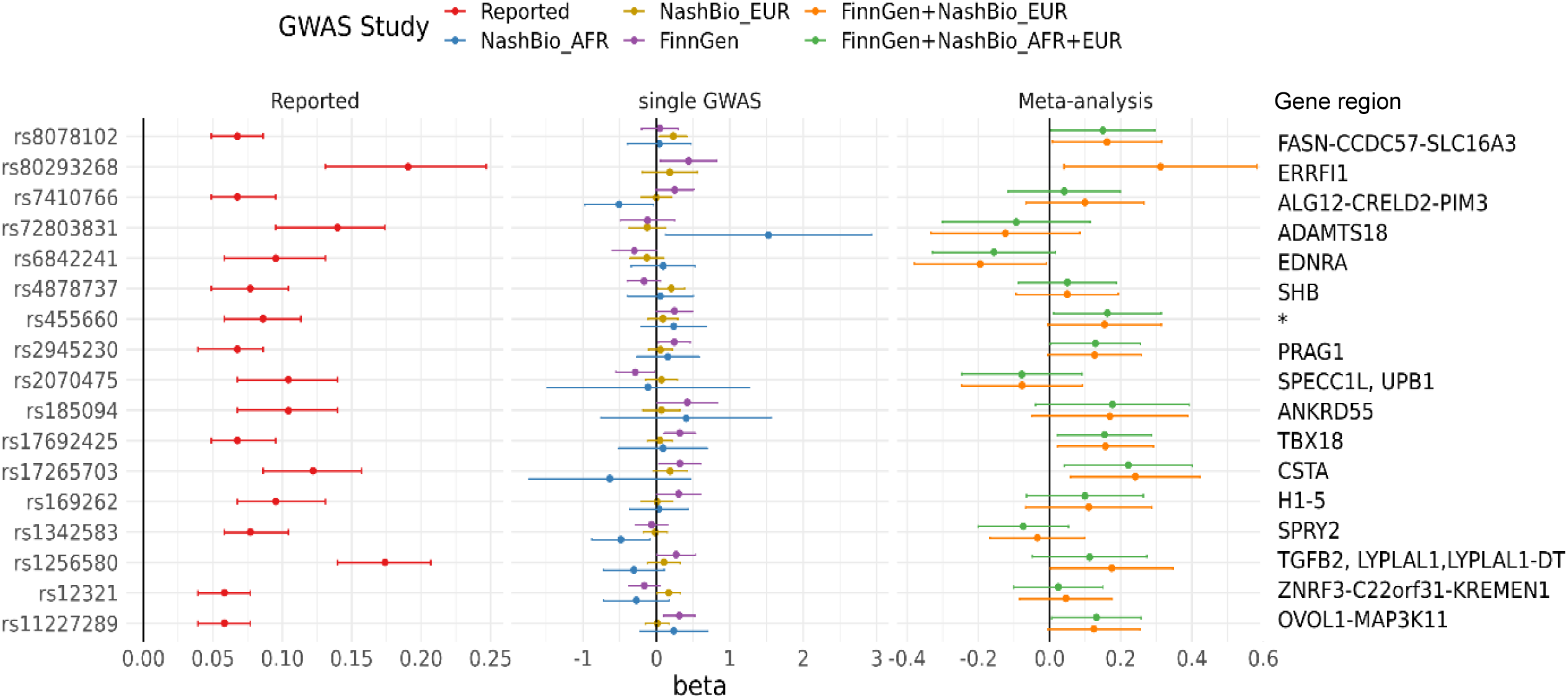
SNP associations that passed nominal significance level in any of the 5 severe acne GWASs at previously reported leading risk variant at acne risk *loci*.

### Polygenic risk score analysis

We evaluated if a polygenic risk score (PRS) constructed with reported lead risk variants of acne at genome-wide significant *loci* and weights also associate with acne severity. The PRS score is statistically significantly associated with severe acne in FinnGen population (OR = 1.77, 95% CI: 1.37 - 2.29, P-value = 1.4x10^-5^), but not associated in EA (OR = 1.22, 95% CI: 0.97 - 1.52, P-value = 0.08) or AA (P-value = 0.99) population in BioVU (Table 4, Figure 3).

**Figure 3.**
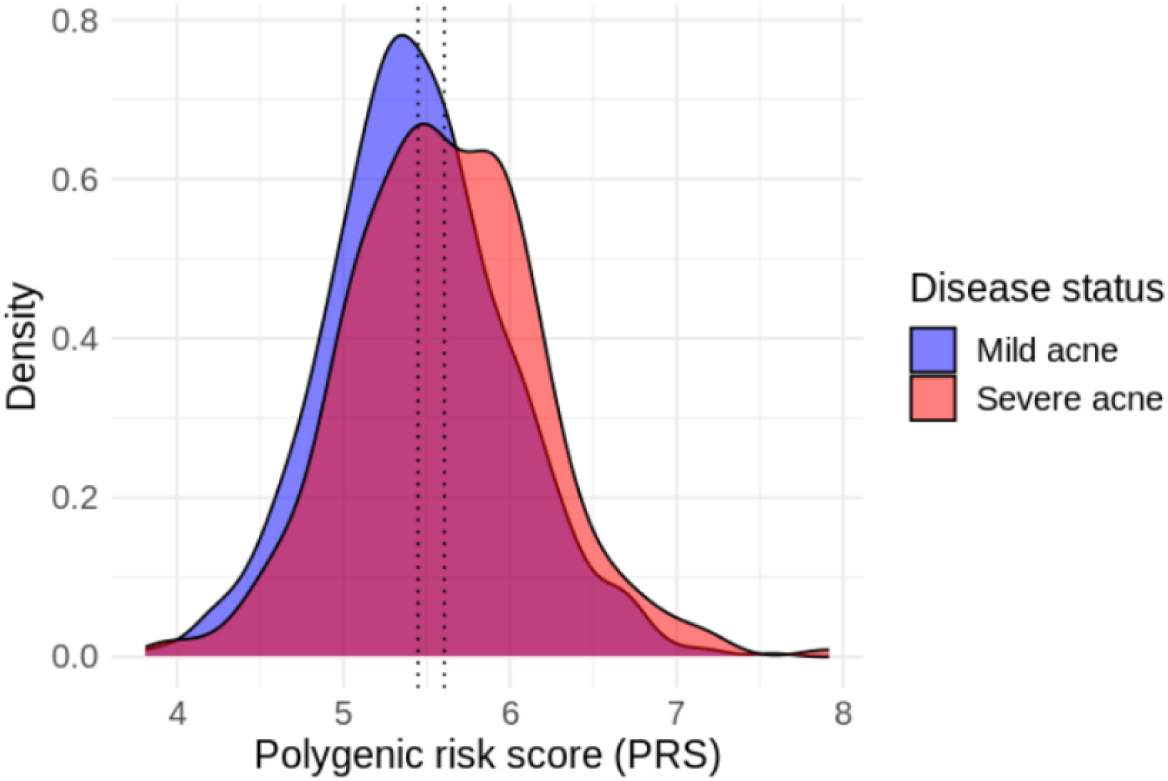
Distribution of acne PRS in FinnGen individuals with severe (red color) or mild (blue color) acne. Dotted vertical lines represent average values of PRS among each group.

**Table 4:** Associations between PRS and severe acne risk by population.

| Study | Mean PRS in cases | Mean PRS in controls | OR (95%CI) | P-value |
| --- | --- | --- | --- | --- |
| FinnGen | 5.61 | 5.45 | 1.77 (1.37 - 2.29) | $1.40 \times 10^{-5}$ |
| BioVU EA | 5.24 | 5.19 | 1.22 (0.97 - 1.52) | $8.40 \times 10^{-2}$ |
| BioVU AA | 5.28 | 5.29 | 1.00 (0.52 - 1.91) | 0.99 |
| BioVU EA + FinnGen | | | 1.46 (1.01 - 2.11) | $4.37 \times 10^{-2}$ |
| BioVU EA + AA + FinnGen | | | 1.37 (1.00 - 1.88) | $4.85 \times 10^{-2}$ |

## Discussion

In this study, we identified one novel genome-wide significant locus associated with severe acne risk among African ancestry population in the BioVU cohort. Leveraging gene-level MR and colocalization analyses, we prioritized six novel genes that confer genetic risks for acne severity. We also identified four pathways that are enriched in severe acne genetic risk using gene-set analysis. Furthermore, we showed that the vast majority of the reported risk alleles for acne onset could not capture genetic risk for acne severity; however, we observed that a PRS constructed by aggregation of these risk variants has significant positive association with acne severity among European ancestry population but not in African ancestry population, possibly due to lower statistical power or lack of portability of PRS across populations^11,12^.

The genome-wide significant SNP rs115325598 is located at the intron region of gene *ATXN7L1*. *ATXN7L1* is poorly characterized functionally. An important paralog of this gene is *ATXN7*, which acts as component of the STAGA transcription coactivator-HAT complex, mediates the interaction of STAGA complex with the CRX and is involved in CRX-dependent gene activation^13,14^, and associates with microtubules and stabilizes the cytoskeletal network^15^. In our integrated differential gene expression analysis, this gene is demonstrated to be upregulated in skin tissue of atopic dermatitis patients and downregulated in epidermal keratinocyte cell in skin tissue of psoriasis vulgaris patients. Through phenome-wide search for associated diseases using our internal gene prioritization scheme, we found this gene has high evidence of association with type 2 diabetes. This is in consistent with previous studies which have shown that both acne vulgaris and type 2 diabetes mellitus are members of the family of mTORC1-driven metabolic diseases^16^. We identified another gene, *ADAM*23, associated with acne severity through our MR and colocalization analysis, and upregulated in keratinocytes in psoriasis patients. *ADAM*23 is a non-catalytic metalloprotease-like protein involved in cell-cell and cell-matrix interaction. This finding highlights the role of skin barrier dysfunction in acne pathophysiology^17^, not only in term of disease onset but also as a factor driving disease severity and progression.

Our study only replicated a small proportion of the known acne susceptibility *loci* at the nominal significance level. The disparity may reflect differences in the biological mechanisms involved in acne onset and acne severity, but could also be influenced by the population-specific genetic determinants, case and control recruitment definitions, and lack of sufficient statistical power. Of note, the polygenic risk score which comprised of aggregated effects of all risk variants is demonstrated to be able to stratify severe versus mild acne, with each one unit increase in PRS is associated with a 1.77- and 1.22-fold elevated risk of developing severe acne. This observation is in concordance of the previous study which showed that the acne PRS has the greatest predictive ability in individuals with severe acne^6^, which implicates the potential application of using a common acne PRS to identify population with high risk of developing severe acne.

One limitation of our study is the small sample size, which makes it underpowered for genome-wide level discovery and replication of the known risk alleles, especially for less common variants with small effect size. Nevertheless, our analysis represents the first study to evaluate the association between genetic variants and acne severity among African ancestry populations and enabled a discovery of a novel genome-wide risk variant. The second limitation of this study is that we defined severe acne cases and controls based on EHR data including ICD codes, frequency of clinical visit, and medication records, rather than using a global acne grading system score (GAGS)^18^, which may cause some selection bias and may impact on the ability to detect true associations. Another limitation is the absence of replication in an independent cohort and functional validation. Future studies are needed to gather more resources to validate our findings.

In summary, our study shows that genetic risk *loci* driving acne onset could not effectively capture genetic risk of acne severity. Disease progression GWAS could contribute to unveiling novel biological mechanisms and may shed light on the discovery of potential therapeutic targets.

## Methods

### Study sample and GWAS

The study participants were from two independent biobanks: the BioVU biorepository extracted by Nashville Biosciences (NashBio), a subsidiary of the Vanderbilt University Medical Center (VUMC) and FinnGen (release 10).

The BioVU has been described previously^19^. Individuals included in this study were BioVU subjects with non-compromised DNA samples available and were previously genotyped on the Illumina Infinium Multiethnic Genotyping Array (MEGAEX) platform. We first selected all subjects with one or more instances of ICD-9/ICD-10 code for acne (L70.0, 706.1) in their electronic medical records (EHR). Among them, severe acne cases were defined as individuals who meet at least one of the following criteria: 1) subjects with a record of isotretinoin, 2) subjects with five or more instances of ICD-9/ICD-10 code of acne in their EHR, or 3) subjects with evidence of treatment with antibiotics for a duration of more than 60 days. Individuals with non-severe acne records and with similar ethnicities were selected as controls. After quality control (QC), a total of 522 severe acne cases, including 449 individuals of European ancestry (EA) and 73 individuals of African ancestry (AA), and 1,214 non-severe acne controls (970 EA individuals and 244 AA individuals) were included in the analysis. Per-allele odds ratios (ORs) and P-value of each variant were estimated using unconditional logistic regression, adjusted for age, sex, and the first 10 principal components (PC). A detailed description of the FinnGen biobank has been published previously^20^. Severe acne cases were defined as individuals who is elder than five years old, with at least five instances of acne diagnosis records (defined by 7061A, 70610, L70.0, L12_ACNE) in their EHR, or with at least five isotretinoin treatment records. Non-severe acne controls were defined as individuals who is elder than five years old, with one to four instances of acne diagnosis in their EHR and with at most four isotretinoin treatment records. After QC, a total of 290 severe acne cases and 908 non-severe acne controls were included in the analysis. Logistic regressions were first performed to determine associated genotyping batches with severe acne. GWAS analysis was then performed using the REGENIE pipeline in FinnGen sandbox^21^, adjusting for age, sex, the first 10 PCs, and the identified associated genotyping batches.

Fixed effect meta-analyses with inverse variance weights were performed using the METAL software^22^ to combine GWAS results from BioVU and FinnGen. Meta-analyses were performed both for EA population only (BioVU EA with FinnGen) and multi-ethnic population (BioVU EA, AA with FinnGen).

### Post-GWAS analyses

#### Variant annotation

We annotated the impact of all variants with *P*<1x10^-6^ using the variant effect predictor (VEP v102)^23^ with the following options: --everything --offline --check_existing. Coding variants were defined as those impacting protein coding transcript annotated as missense variant or predicted to have “high” impact. We also retrieved predicted gain or loss of function variants^6^. We also mapped variants to genes using activity-by-contact (ABC) enhancer-promoter interactions^10^. We used “ABCmax” to represent variant-gene pairs with the highest ABC score.

#### Mendelian randomization and colocalization

We conducted mendelian randomization (MR) using the TwoSampleMR R package^24^. The following exposures were considered: protein quantitative trait loci (pQTL) from Sun et al^25^, and expression quantitative trait loci from Blueprint^26^, eQTLGen^27^ and other datasets from the EBI eQTL catalogue ^26–41^. For each of those studies, variants with a MAF < 1% were excluded. Variants were clumped using PLINK2^42^ using the options – clump-p1 1 –clump-p2 1 –clump-r2 0.01 – clump-kb 10000. Only genes 250kb around significant *loci* were considered in this analysis. For each QTL, independent variants with *P* < 1x10^-4^ were used as instruments. When more than one instrument was present, the inverse variant weighted approach was used, otherwise the Wald Ratio approach was used. False Discovery Rate (FDR) corrections were applied to adjust *P-values* in multiple testing, and a threshold of FDR < 0.05 was used to determine significance. For genes with significant MR results, we performed colocalization analyses to detect shared causal SNPs between molecular-QTL and acne severity using the coloc R package with default priors^43^, using a region of 250kb around the local lead GWAS variant. We set a threshold of colocalization posterior probability at H4 (PP.H4) ≥ 0.8, and genes with both significant MR and colocalization were considered as potential targeted molecular. Only autosomes were included in this analysis.

#### Gene-set analyses using MAGMA

We performed gene-set level analyses using MAGMA (v1.10)^44^. SNP annotation to genes was done using the 1000 Genomes Phase 3 European panel and GRCh38 (downloaded from http://ctglab.nl/software/magma). Gene sets were downloaded from the Molecular Signatures Database (MSigDB) ^45,46^, including 7763 gene sets in Gene Ontology (GO), 186 in Kyoto Encyclopedia of Genes and Genomes (KEGG), 292 in BioCarta, 1,635 in Reactome and 4,872 in ImmuneSigDB.

### Associations of identified genes with other phenotypes

For each severe acne related gene identified from GWAS (nearest gene of the genome-wide significant risk variant), MR and Colocalization, we performed a phenome-wide search to identify other diseases that showed evidence of association with the focal severe acne-related gene. We employed an in-house gene prioritization scheme, which contained precalculated results across 6,918 GWAS from 4 sources, including UK Biobank, FinnGen release 10 (R10), EBI GWAS catalog, and meta-analyses of UK Biobank, FinnGen R10, and Estonian biobank [ref: GiTIPs paper]. Briefly, this gene-ranking scheme used a combinatory approach to prioritize disease putative genes based on MR, eQTL and pQTL colocalization, ABC enhancer-promoter interactions, and variant annotations. This approach mitigates the pleiotropic effect of eQTL while retaining important information about directionality and has been shown to be enriched for “gold standard” genes and drug targets with successful clinical trial [ref: GiTIPs paper].

### Differential expression in other diseases

We further examined whether the identified acne severity related genes are differentially expressed in diseased compared to normal tissues of other diseases. Differential gene expressions (DGE) were extracted from integrated transcriptomics service from Qiagen Omicsoft database (QIAGEN OmicSoft DiseaseLand^47^). We extracted both microarray and RNA-seq based transcriptomic (TxP) studies and disease comparisons/contrasts available for all studies in the DiseaseLand in the immunology and inflammation, neurodegenerative and metabolic diseases^48^. Common ontologies were applied to all samples, tissues, diseases, treatments, and comparison types to systematize database curation. Database raw data was derived from published repositories including gene expression omnibus (GEO) and ArrayExpress (https://www.ebi.ac.uk/biostudies/arrayexpress). In total, the integrated database consists of >250,000 samples with TxP profiles, 32,000 study comparison contrasts in ∼ 700 disease areas. The contrasts that were of interest to us are “Disease.vs.Normal” which we subsequently used to perform meta-analysis.

Several studies have delved into the exploration of integrated transcriptomics data through various meta-analyses ^48,49^. We opted for the Fisher’s exact test for its straightforward implementation, heightened sensitivity to small sample sizes, non-parametric nature, and platform independence across transcriptomics techniques (such as microarray and RNA-seq). Overall, meta-analysis examined whether a gene_i is a recurrent DGE enriched for a given disease, tissue, or sample (e.g.: Alzheimer’s disease and brain tissue) when compared to recurrency of the gene in the entire database. We constructed a contingency table as outlined in the methodology suggested earlier by Wang et al.^50^ where we used a cut-off of adjusted P values (*P-adj*<0.05) & abs(log2Fold-change)>1.0 to define a DGE, and at least 70% of studies having the same direction of log2Fold-Change (positive or negative). A recurrent DGE enriched across different studies increases the confidence of the gene in playing a role in the disease mechanism of action (MoA).

### Gene expression levels by cell types and tissues

We then assessed the expression levels of the identified acne severity related genes by tissues and cell types. Baseline gene expression data were retrieved from GTEx and Expression Atlas (http://www.ebi.ac.uk/gxa) ^51^, and single-cell transcriptomics were obtained from the BioTuring Single-cell gene expression database^52^.

### Association testing of known risk regions

We examined whether the previously reported genetic risk *loci* of acne also contribute to severe acne risk. Directional consistency of effect was defined as ORs in the current GWASs that were in the same direction of effect (ie, >1) as those reported in the discovery population. A nominal P value of .05 was used to determine statistical significance.

### Polygenic risk score analysis

The aggregate effect of known risk alleles was examined using a weighted polygenic risk score (PRS) for each individual: 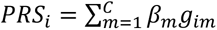, where *g_im_* is the risk allele dosage for individual i at SNP m, C defines risk SNPs at 66 known acne susceptibility *loci*, and *β_m_* is the weight for SNP m. The association between PRS and severe acne risk was examined separately in each dataset using logistic regression, adjusting for the first 10 PCs, age, and sex. The results were combined using a fixed-effect meta-analysis with inverse variance weights.

## ETHICS APPROVAL AND CONSENT TO PARTICIPATE

Patients and control subjects in FinnGen provided informed consent for biobank research, based on the Finnish Biobank Act. Alternatively, separate research cohorts, collected prior the Finnish Biobank Act came into effect (in September 2013) and start of FinnGen (August 2017), were collected based on study-specific consents and later transferred to the Finnish biobanks after approval by Fimea, the National Supervisory Authority for Welfare and Health. Recruitment protocols followed the biobank protocols approved by Fimea. The Coordinating Ethics Committee of the Hospital District of Helsinki and Uusimaa (HUS) approved the FinnGen study protocol Nr HUS/990/2017. The FinnGen study is approved by Finnish Institute for Health and Welfare (permit numbers: THL/2031/6.02.00/2017, THL/1101/5.05.00/2017, THL/341/6.02.00/2018, THL/2222/6.02.00/2018, THL/283/6.02.00/2019, THL/1721/5.05.00/2019, THL/1524/5.05.00/2020, and THL/2364/14.02/2020), Digital and population data service agency (permit numbers: VRK43431/2017-3, VRK/6909/2018-3, VRK/4415/2019-3), the Social Insurance Institution (permit numbers: KELA 58/522/2017, KELA 131/522/2018, KELA 70/522/2019, KELA 98/522/2019, KELA 138/522/2019, KELA 2/522/2020, KELA 16/522/2020 and Statistics Finland (permit numbers: TK-53-1041-17 and TK-53-90-20). The Biobank Access Decisions for FinnGen samples and data utilized in FinnGen Data Freeze 6 include: THL Biobank BB2017_55, BB2017_111, BB2018_19, BB_2018_34, BB_2018_67, BB2018_71, BB2019_7, BB2019_8, BB2019_26, BB2020_1, Finnish Red Cross Blood Service Biobank 7.12.2017, Helsinki Biobank HUS/359/2017, Auria Biobank AB17-5154, Biobank Borealis of Northern Finland_2017_1013, Biobank of Eastern Finland 1186/2018, Finnish Clinical Biobank Tampere MH0004, Central Finland Biobank 1-2017, and Terveystalo Biobank STB 2018001. UK Biobank has received ethical approval from the NHS National Research Ethics Service North West (approval numbers 11/NW/0382 and 16/NW/0274). All participants provided written informed consent.

## DATA AVAILABILITY

All data produced in the present study are available upon reasonable request to the authors.

## ACKNOWLEDGEMENT

FinnGen

We want to acknowledge the participants and investigators of FinnGen study. The FinnGen project is funded by two grants from Business Finland (HUS 4685/31/2016 and UH 4386/31/2016) and the following industry partners: AbbVie Inc., AstraZeneca UK Ltd, Biogen MA Inc., Bristol Myers Squibb (and Celgene Corporation & Celgene International II Sàrl), Genentech Inc., Merck Sharp & Dohme LCC, Pfizer Inc., GlaxoSmithKline Intellectual Property Development Ltd., Sanofi US Services Inc., Maze Therapeutics Inc., Janssen Biotech Inc, Novartis Pharma AG, and Boehringer Ingelheim International GmbH. Following biobanks are acknowledged for delivering biobank samples to FinnGen: Auria Biobank (www.auria.fi/biopankki), THL Biobank (www.thl.fi/biobank), Helsinki Biobank (www.helsinginbiopankki.fi), Biobank Borealis of Northern Finland (https://www.ppshp.fi/Tutkimus-ja-opetus/Biopankki/Pages/Biobank-Borealis-briefly-in-English.aspx), Finnish Clinical Biobank Tampere (www.tays.fi/en-US/Research_and_development/Finnish_Clinical_Biobank_Tampere), Biobank of Eastern Finland (www.ita-suomenbiopankki.fi/en), Central Finland Biobank (www.ksshp.fi/fi-FI/Potilaalle/Biopankki), Finnish Red Cross Blood Service Biobank (www.veripalvelu.fi/verenluovutus/biopankkitoiminta), Terveystalo Biobank (www.terveystalo.com/fi/Yritystietoa/Terveystalo-Biopankki/Biopankki/) and Arctic Biobank (https://www.oulu.fi/en/university/faculties-and-units/faculty-medicine/northern-finland-birth-cohorts-and-arctic-biobank). All Finnish Biobanks are members of BBMRI.fi infrastructure (www.bbmri.fi). Finnish Biobank Cooperative -FINBB (https://finbb.fi/) is the coordinator of BBMRI-ERIC operations in Finland. The Finnish biobank data can be accessed through the Fingenious® services (https://site.fingenious.fi/en/) managed by FINBB.

BioVU

The datasets used for the analyses described were obtained from Vanderbilt University Medical Center’s BioVU, which are supported by numerous sources: institutional funding, private agencies, and federal grants. These include NIH funded Shared Instrumentation Grant S10OD017985, S10RR025141, and S10OD025092; CTSA grants UL1TR002243, UL1TR000445, and UL1RR024975. Genomic data are also supported by investigator-led projects that include U01HG004798, R01NS032830, RC2GM092618, P50GM115305, U01HG006378, U19HL065962, and R01HD074711.

UK Biobank

This research has been conducted using the UK Biobank Resource (www.ukbiobank.ac.uk), a large-scale biomedical database and research resource containing genetic, lifestyle and health information from 500,000 UK participants. UK Biobank is supported by its founding funders the Wellcome Trust and UK Medical Research Council, as well as the Department of Health, Scottish Government, the Northwest Regional Development Agency, British Heart Foundation and Cancer Research UK. The UK biobank pan-ancestry analysis was conducted under project ID 31063 (https://pan.ukbb.broadinstitute.org).

Estonian Biobank

Estonian Biobank research was supported by the European Union through Horizon 2020 research and innovation programme under grant no 810645 and through the European Regional Development Fund project no. MOBEC008, by the Estonian Research Council grant PUT (PRG1291, PRG687 and PRG184) and by the European Union through the European Regional Development Fund project no. MOBERA21 (ERA-CVD project DETECT ARRHYTHMIAS, GA no JTC2018-009), Project No. 2014-2020.4.01.15-0012 and Project No. 2014-2020.4.01.16-0125.

