## Supplementary figure 1-2 for "Genome-wide association study analysis of disease severity in Acne reveals novel biological insights"

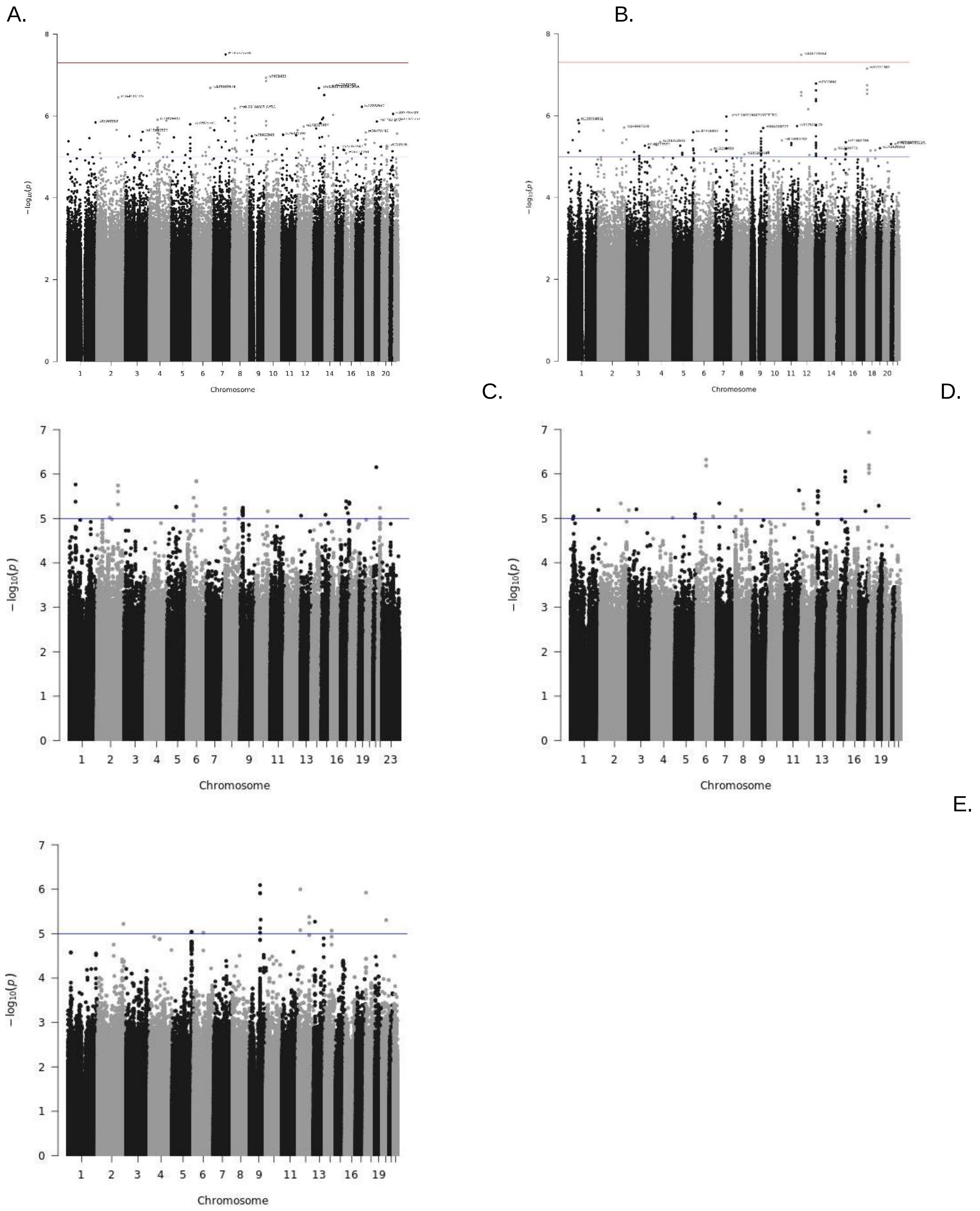
A.
B.
C.
D.
E.
$-\log_{10}(p)$
Chromosome

Supplementary Figure 1: Manhattan plot of Genome-wide association studies (GWASs) of severe acne in A) African ancestry (AA) population in BioVU cohort, B) European ancestry (EA) population in BioVU cohort, C) FinnGen Release 10 (R10), D) meta-analysis of BioVU EA population and FinnGen R10 and E) meta-analysis of BioVU EA, AA population and FinnGen R10. Manhattan plot shows all genotyped and imputed results, excluding SNPs with MAF $< 0.01$ in severe acne cases and imputation score $< 0.8$. x-Axis is chromosome position, y-axis is $-\log10$(p-value). The red line represents the genome-wide significant cut-off value of $p = 5 \times 10^{-8}$, and blue line represents suggestive significance level at $1 \times 10^{-5}$.

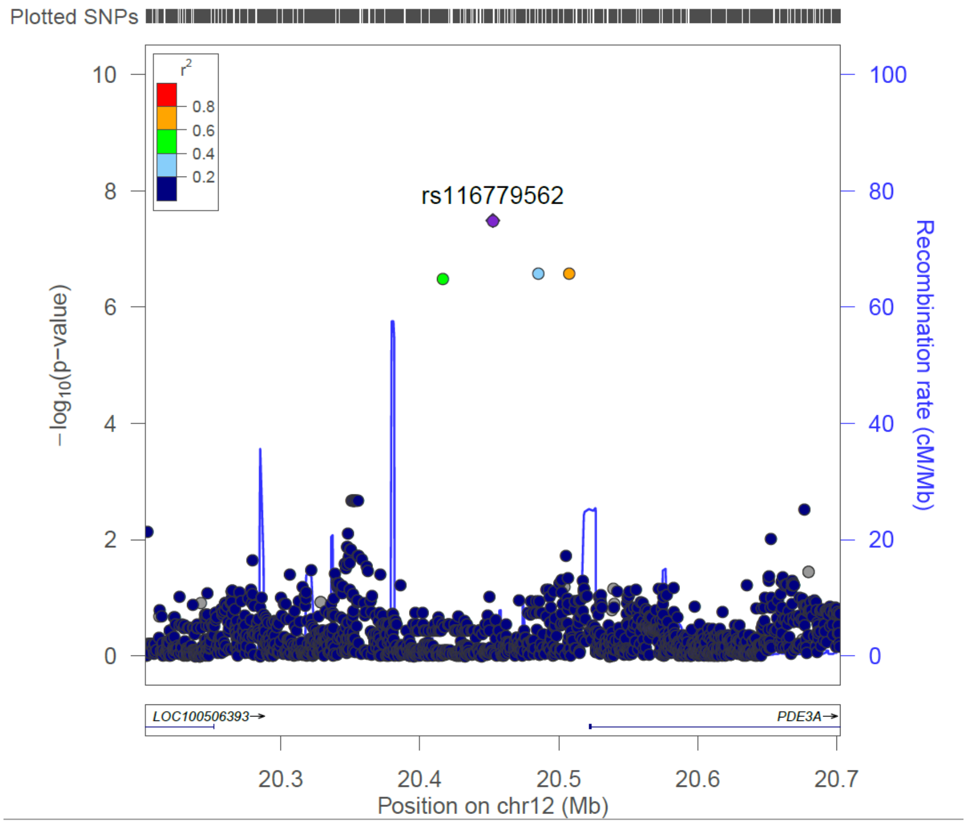


Supplementary figure 2: Regional association plots of the 12p12.2 risk region for acne severity among European ancestry (EA) population in BioVU cohort. Single-nucleotide polymorphisms (SNPs) are plotted by position (x-axis) and −log10 p-value (y-axis). $r^2$ was estimated from EA individuals in phase III 1000 Genomes Project (1KGP) data. The most statistically significant associated SNP (purple diamond) at 12p12.2 is rs116779562. The surrounding SNPs are colored to indicate pairwise correlation with the index SNP.
