## Supplementary figure 3 for "Genome-wide association study analysis of disease severity in Acne reveals novel biological insights"

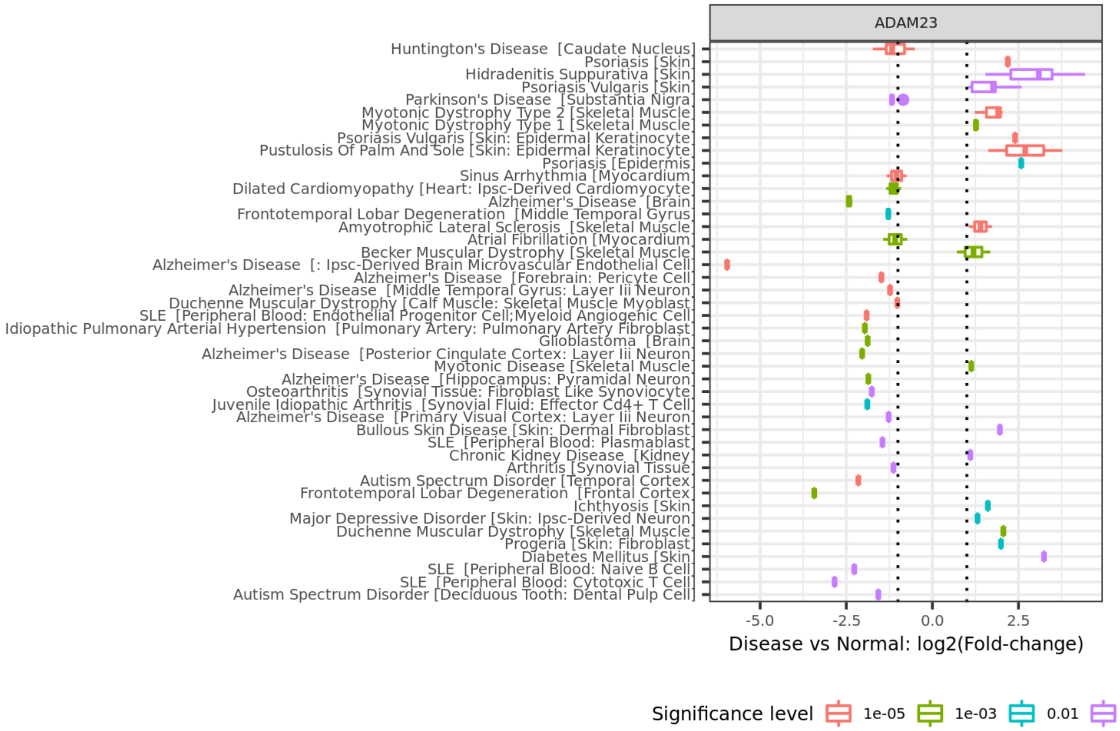

ADAM23
Huntington's Disease [Caudate Nucleus]
Psoriasis [Skin]
Hidradenitis Suppurativa [Skin]
Psoriasis Vulgaris [Skin]
Parkinson's Disease [Substantia Nigra]
Myotonic Dystrophy Type 2 [Skeletal Muscle]
Myotonic Dystrophy Type 1 [Skeletal Muscle]
Psoriasis Vulgaris [Skin: Epidermal Keratinocyte]
Pustulosis Of Palm And Sole [Skin: Epidermal Keratinocyte]
Psoriasis [Epidermis]
Sinus Arrhythmia [Myocardium]
Dilated Cardiomyopathy [Heart: Ipsc-Derived Cardiomyocyte]
Alzheimer's Disease [Brain]
Frontotemporal Lobar Degeneration [Middle Temporal Gyrus]
Amyotrophic Lateral Sclerosis [Skeletal Muscle]
Atrial Fibrillation [Myocardium]
Becker Muscular Dystrophy [Skeletal Muscle]
Alzheimer's Disease [: Ipsc-Derived Brain Microvascular Endothelial Cell]
Alzheimer's Disease [Forebrain: Pericyte Cell]
Alzheimer's Disease [Middle Temporal Gyrus: Layer Iii Neuron]
Duchenne Muscular Dystrophy [Calf Muscle: Skeletal Muscle Myoblast]
SLE [Peripheral Blood: Endothelial Progenitor Cell;Myeloid Angiogenic Cell]
Idiopathic Pulmonary Arterial Hypertension [Pulmonary Artery: Pulmonary Artery Fibroblast]
Glioblastoma [Brain]
Alzheimer's Disease [Posterior Cingulate Cortex: Layer Iii Neuron]
Myotonic Disease [Skeletal Muscle]
Alzheimer's Disease [Hippocampus: Pyramidal Neuron]
Osteoarthritis [Synovial Tissue: Fibroblast Like Synoviocyte]
Juvenile Idiopathic Arthritis [Synovial Fluid: Effector Cd4+ T Cell]
Alzheimer's Disease [Primary Visual Cortex: Layer Iii Neuron]
Bullous Skin Disease [Skin: Dermal Fibroblast]
SLE [Peripheral Blood: Plasmablast]
Chronic Kidney Disease [Kidney]
Arthritis [Synovial Tissue]
Autism Spectrum Disorder [Temporal Cortex]
Frontotemporal Lobar Degeneration [Frontal Cortex]
Ichthyosis [Skin]
Major Depressive Disorder [Skin: Ipsc-Derived Neuron]
Duchenne Muscular Dystrophy [Skeletal Muscle]
Progeria [Skin: Fibroblast]
Diabetes Mellitus [Skin]
SLE [Peripheral Blood: Naive B Cell]
SLE [Peripheral Blood: Cytotoxic T Cell]
Autism Spectrum Disorder [Deciduous Tooth: Dental Pulp Cell]
-5.0
-2.5
0.0
2.5
Disease vs Normal: log2(Fold-change)
Significance level
1e-05
1e-03
0.01


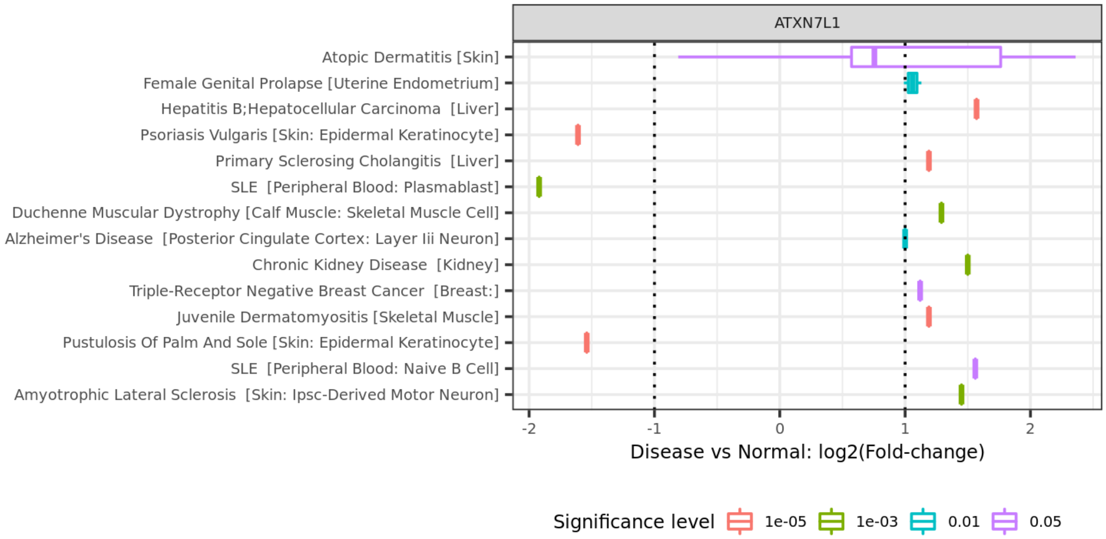

ATXN7L1
Atopic Dermatitis [Skin]
Female Genital Prolapse [Uterine Endometrium]
Hepatitis B;Hepatocellular Carcinoma [Liver]
Psoriasis Vulgaris [Skin: Epidermal Keratinocyte]
Primary Sclerosing Cholangitis [Liver]
SLE [Peripheral Blood: Plasmablast]
Duchenne Muscular Dystrophy [Calf Muscle: Skeletal Muscle Cell]
Alzheimer's Disease [Posterior Cingulate Cortex: Layer Iii Neuron]
Chronic Kidney Disease [Kidney]
Triple-Receptor Negative Breast Cancer [Breast:]
Juvenile Dermatomyositis [Skeletal Muscle]
Pustulosis Of Palm And Sole [Skin: Epidermal Keratinocyte]
SLE [Peripheral Blood: Naive B Cell]
Amyotrophic Lateral Sclerosis [Skin: Ipsc-Derived Motor Neuron]
-2
-1
0
1
2
Disease vs Normal: log2(Fold-change)
Significance level
1e-05
1e-03
0.01
0.05

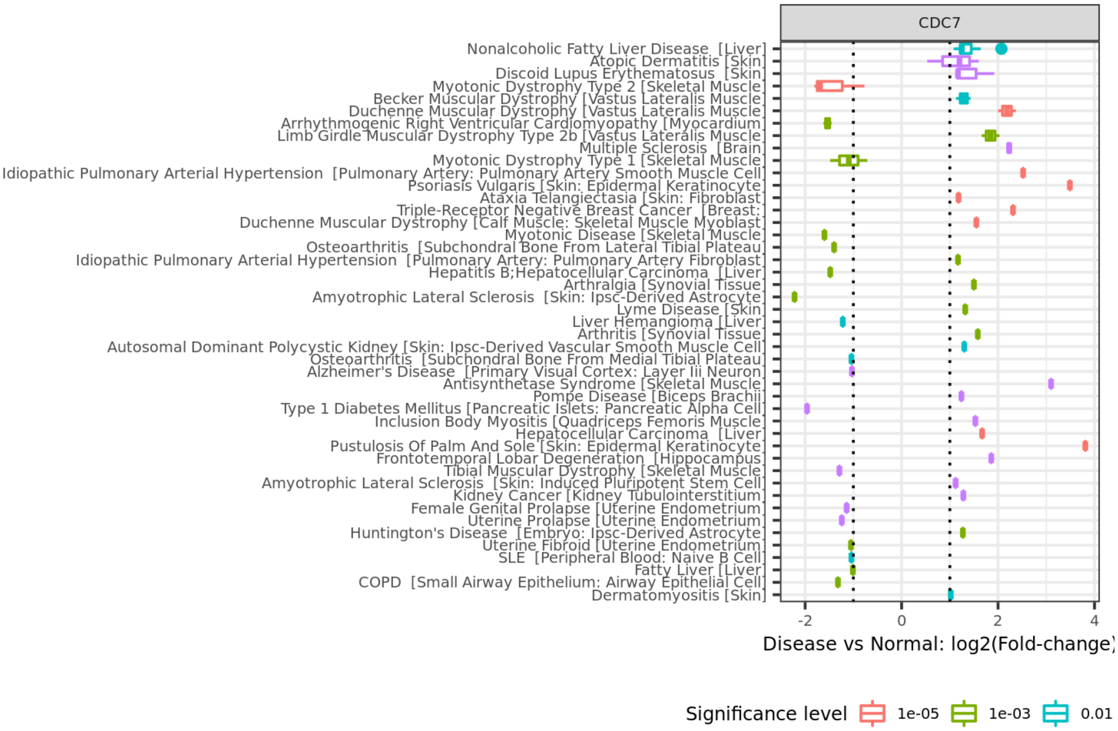
CDC7
Nonalcoholic Fatty Liver Disease [Liver]
Atopic Dermatitis [Skin]
Discoid Lupus Erythematosus [Skin]
Myotonic Dystrophy Type 2 [Skeletal Muscle]
Becker Muscular Dystrophy [Vastus Lateralis Muscle]
Duchenne Muscular Dystrophy [Vastus Lateralis Muscle]
Arrhythmogenic Right Ventricular Cardiomyopathy [Myocardium]
Limb Girdle Muscular Dystrophy Type 2b [Vastus Lateralis Muscle]
Multiple Sclerosis [Brain]
Myotonic Dystrophy Type 1 [Skeletal Muscle]
Idiopathic Pulmonary Arterial Hypertension [Pulmonary Artery: Pulmonary Artery Smooth Muscle Cell]
Psoriasis Vulgaris [Skin: Epidermal Keratinocyte]
Ataxia Telangiectasia [Skin: Fibroblast]
Triple-Receptor Negative Breast Cancer [Breast]
Duchenne Muscular Dystrophy [Calf Muscle: Skeletal Muscle Myoblast]
Myotonic Disease [Skeletal Muscle]
Osteoarthritis [Subchondral Bone From Lateral Tibial Plateau]
Idiopathic Pulmonary Arterial Hypertension [Pulmonary Artery: Pulmonary Artery Fibroblast]
Hepatitis B;Hepatocellular Carcinoma [Liver]
Arthralgia [Synovial Tissue]
Amyotrophic Lateral Sclerosis [Skin: Ipsc-Derived Astrocyte]
Lyme Disease [Skin]
Liver Hemangioma [Liver]
Arthritis [Synovial Tissue]
Autosomal Dominant Polycystic Kidney [Skin: Ipsc-Derived Vascular Smooth Muscle Cell]
Osteoarthritis [Subchondral Bone From Medial Tibial Plateau]
Alzheimer's Disease [Primary Visual Cortex: Layer Iii Neuron]
Antisynthetase Syndrome [Skeletal Muscle]
Pompe Disease [Biceps Brachii]
Type 1 Diabetes Mellitus [Pancreatic Islets: Pancreatic Alpha Cell]
Inclusion Body Myositis [Quadriceps Femoris Muscle]
Hepatocellular Carcinoma [Liver]
Pustulosis Of Palm And Sole [Skin: Epidermal Keratinocyte]
Frontotemporal Lobar Degeneration [Hippocampus]
Tibial Muscular Dystrophy [Skeletal Muscle]
Amyotrophic Lateral Sclerosis [Skin: Induced Pluripotent Stem Cell]
Kidney Cancer [Kidney Tubulointerstitium]
Female Genital Prolapse [Uterine Endometrium]
Uterine Prolapse [Uterine Endometrium]
Huntington's Disease [Embryo: Ipsc-Derived Astrocyte]
Uterine Fibroid [Uterine Endometrium]
SLE [Peripheral Blood: Naive B Cell]
Fatty Liver [Liver]
COPD [Small Airway Epithelium: Airway Epithelial Cell]
Dermatomyositis [Skin]
-2
0
2
4
Disease vs Normal: log2(Fold-change)
Significance level
1e-05
1e-03
0.01

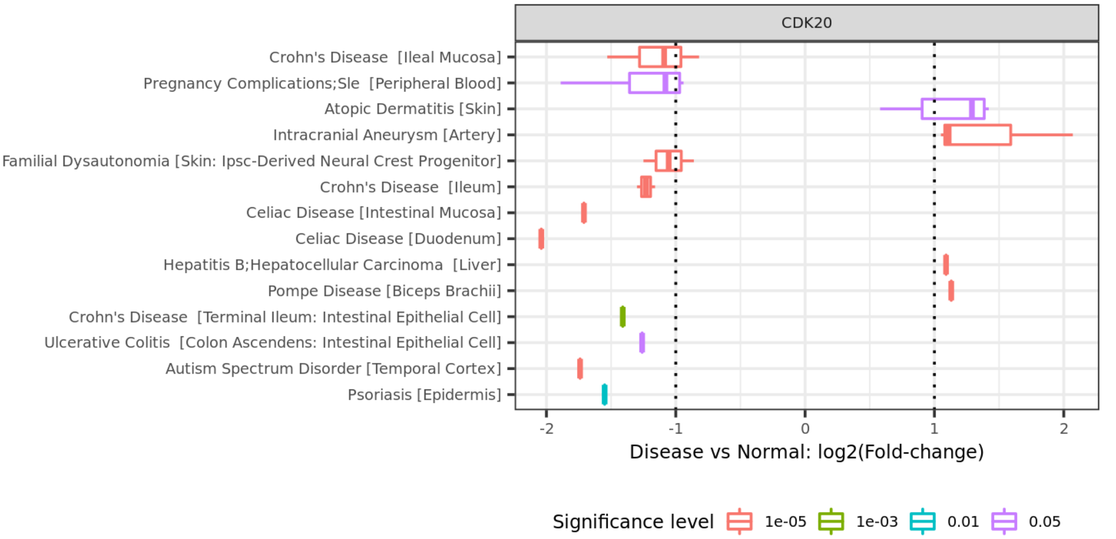
CDK20
Crohn's Disease [Ileal Mucosa]
Pregnancy Complications;Sle [Peripheral Blood]
Atopic Dermatitis [Skin]
Intracranial Aneurysm [Artery]
Familial Dysautonomia [Skin: Ipsc-Derived Neural Crest Progenitor]
Crohn's Disease [Ileum]
Celiac Disease [Intestinal Mucosa]
Celiac Disease [Duodenum]
Hepatitis B;Hepatocellular Carcinoma [Liver]
Pompe Disease [Biceps Brachii]
Crohn's Disease [Terminal Ileum: Intestinal Epithelial Cell]
Ulcerative Colitis [Colon Ascendens: Intestinal Epithelial Cell]
Autism Spectrum Disorder [Temporal Cortex]
Psoriasis [Epidermis]
-2
-1
0
1
2
Disease vs Normal: log2(Fold-change)
Significance level
1e-05
1e-03
0.01
0.05

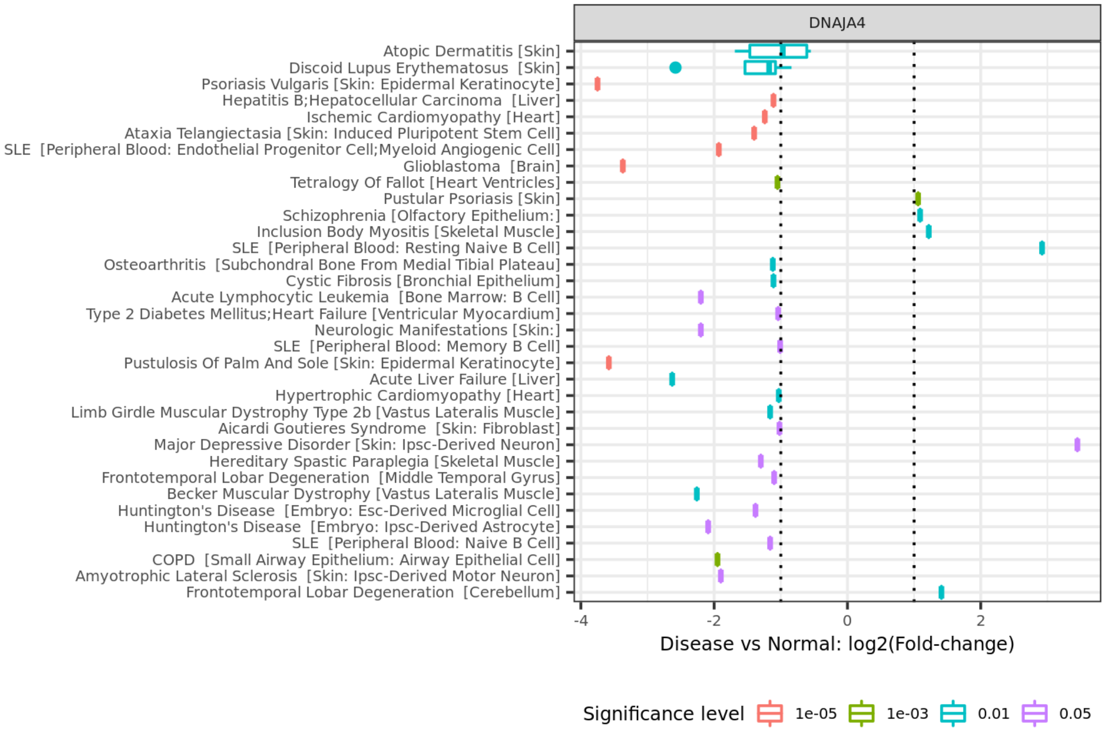
DNAJA4
Atopic Dermatitis [Skin]
Discoid Lupus Erythematosus [Skin]
Psoriasis Vulgaris [Skin: Epidermal Keratinocyte]
Hepatitis B;Hepatocellular Carcinoma [Liver]
Ischemic Cardiomyopathy [Heart]
Ataxia Telangiectasia [Skin: Induced Pluripotent Stem Cell]
SLE [Peripheral Blood: Endothelial Progenitor Cell;Myeloid Angiogenic Cell]
Glioblastoma [Brain]
Tetralogy Of Fallot [Heart Ventricles]
Pustular Psoriasis [Skin]
Schizophrenia [Olfactory Epithelium:]
Inclusion Body Myositis [Skeletal Muscle]
SLE [Peripheral Blood: Resting Naive B Cell]
Osteoarthritis [Subchondral Bone From Medial Tibial Plateau]
Cystic Fibrosis [Bronchial Epithelium]
Acute Lymphocytic Leukemia [Bone Marrow: B Cell]
Type 2 Diabetes Mellitus;Heart Failure [Ventricular Myocardium]
Neurologic Manifestations [Skin:]
SLE [Peripheral Blood: Memory B Cell]
Pustulosis Of Palm And Sole [Skin: Epidermal Keratinocyte]
Acute Liver Failure [Liver]
Hypertrophic Cardiomyopathy [Heart]
Limb Girdle Muscular Dystrophy Type 2b [Vastus Lateralis Muscle]
Aicardi Goutieres Syndrome [Skin: Fibroblast]
Major Depressive Disorder [Skin: Ipsc-Derived Neuron]
Hereditary Spastic Paraplegia [Skeletal Muscle]
Frontotemporal Lobar Degeneration [Middle Temporal Gyrus]
Becker Muscular Dystrophy [Vastus Lateralis Muscle]
Huntington's Disease [Embryo: Esc-Derived Microglial Cell]
Huntington's Disease [Embryo: Ipsc-Derived Astrocyte]
SLE [Peripheral Blood: Naive B Cell]
COPD [Small Airway Epithelium: Airway Epithelial Cell]
Amyotrophic Lateral Sclerosis [Skin: Ipsc-Derived Motor Neuron]
Frontotemporal Lobar Degeneration [Cerebellum]
-4
-2
0
2
Disease vs Normal: log2(Fold-change)
Significance level
1e-05
1e-03
0.01
0.05

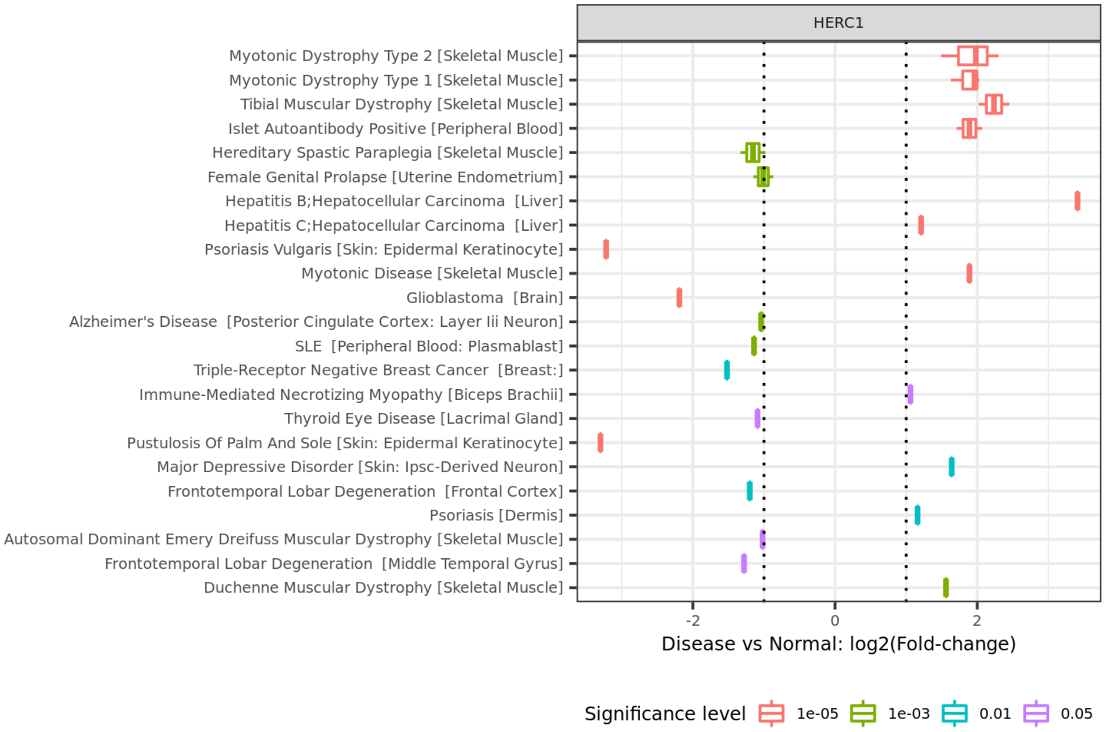
HERC1
Myotonic Dystrophy Type 2 [Skeletal Muscle]
Myotonic Dystrophy Type 1 [Skeletal Muscle]
Tibial Muscular Dystrophy [Skeletal Muscle]
Islet Autoantibody Positive [Peripheral Blood]
Hereditary Spastic Paraplegia [Skeletal Muscle]
Female Genital Prolapse [Uterine Endometrium]
Hepatitis B;Hepatocellular Carcinoma [Liver]
Hepatitis C;Hepatocellular Carcinoma [Liver]
Psoriasis Vulgaris [Skin: Epidermal Keratinocyte]
Myotonic Disease [Skeletal Muscle]
Glioblastoma [Brain]
Alzheimer's Disease [Posterior Cingulate Cortex: Layer Iii Neuron]
SLE [Peripheral Blood: Plasmablast]
Triple-Receptor Negative Breast Cancer [Breast:]
Immune-Mediated Necrotizing Myopathy [Biceps Brachii]
Thyroid Eye Disease [Lacrimal Gland]
Pustulosis Of Palm And Sole [Skin: Epidermal Keratinocyte]
Major Depressive Disorder [Skin: Ipsc-Derived Neuron]
Frontotemporal Lobar Degeneration [Frontal Cortex]
Psoriasis [Dermis]
Autosomal Dominant Emery Dreifuss Muscular Dystrophy [Skeletal Muscle]
Frontotemporal Lobar Degeneration [Middle Temporal Gyrus]
Duchenne Muscular Dystrophy [Skeletal Muscle]
-2
0
2
Disease vs Normal: log2(Fold-change)
Significance level
1e-05
1e-03
0.01
0.05

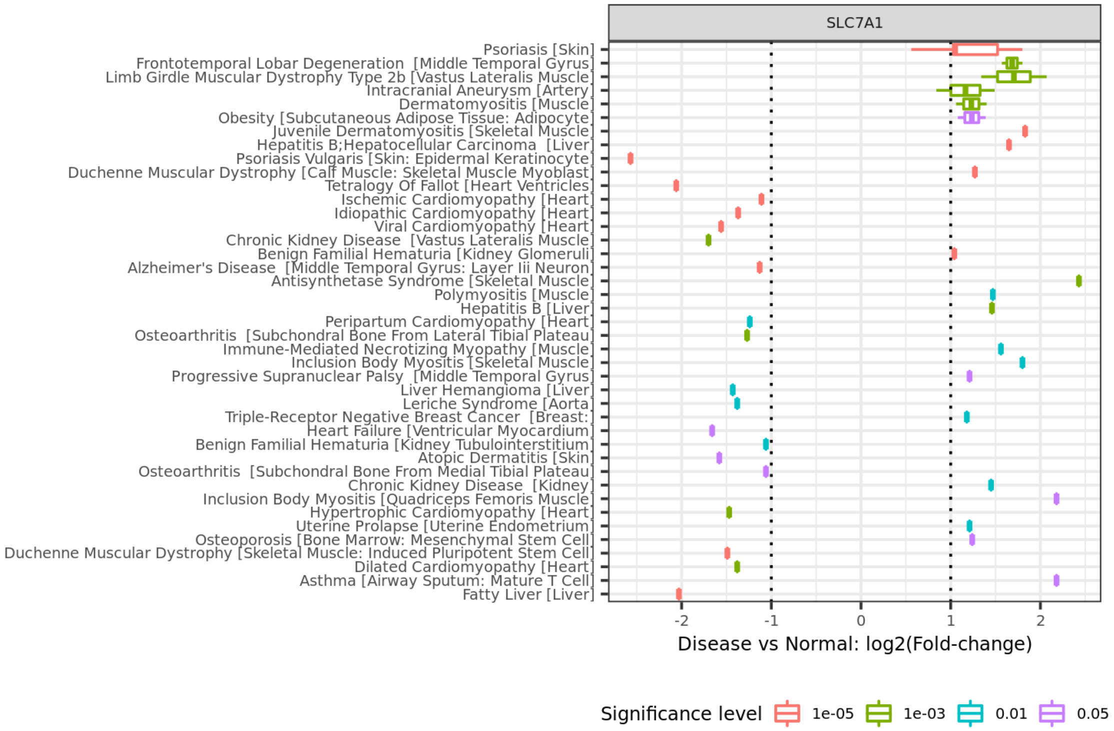
SLC7A1
Psoriasis [Skin]
Frontotemporal Lobar Degeneration [Middle Temporal Gyrus]
Limb Girdle Muscular Dystrophy Type 2b [Vastus Lateralis Muscle]
Intracranial Aneurysm [Artery]
Dermatomyositis [Muscle]
Obesity [Subcutaneous Adipose Tissue: Adipocyte]
Juvenile Dermatomyositis [Skeletal Muscle]
Hepatitis B;Hepatocellular Carcinoma [Liver]
Psoriasis Vulgaris [Skin: Epidermal Keratinocyte]
Duchenne Muscular Dystrophy [Calf Muscle: Skeletal Muscle Myoblast]
Tetralogy Of Fallot [Heart Ventricles]
Ischemic Cardiomyopathy [Heart]
Idiopathic Cardiomyopathy [Heart]
Viral Cardiomyopathy [Heart]
Chronic Kidney Disease [Vastus Lateralis Muscle]
Benign Familial Hematuria [Kidney Glomeruli]
Alzheimer's Disease [Middle Temporal Gyrus: Layer Iii Neuron]
Antisynthetase Syndrome [Skeletal Muscle]
Polymyositis [Muscle]
Hepatitis B [Liver]
Peripartum Cardiomyopathy [Heart]
Osteoarthritis [Subchondral Bone From Lateral Tibial Plateau]
Immune-Mediated Necrotizing Myopathy [Muscle]
Inclusion Body Myositis [Skeletal Muscle]
Progressive Supranuclear Palsy [Middle Temporal Gyrus]
Liver Hemangioma [Liver]
Leriche Syndrome [Aorta]
Triple-Receptor Negative Breast Cancer [Breast:]
Heart Failure [Ventricular Myocardium]
Benign Familial Hematuria [Kidney Tubulointerstitium]
Atopic Dermatitis [Skin]
Osteoarthritis [Subchondral Bone From Medial Tibial Plateau]
Chronic Kidney Disease [Kidney]
Inclusion Body Myositis [Quadriceps Femoris Muscle]
Hypertrophic Cardiomyopathy [Heart]
Uterine Prolapse [Uterine Endometrium]
Osteoporosis [Bone Marrow: Mesenchymal Stem Cell]
Duchenne Muscular Dystrophy [Skeletal Muscle: Induced Pluripotent Stem Cell]
Dilated Cardiomyopathy [Heart]
Asthma [Airway Sputum: Mature T Cell]
Fatty Liver [Liver]
-2
-1
0
1
2
Disease vs Normal: log2(Fold-change)
Significance level
1e-05
1e-03
0.01
0.05

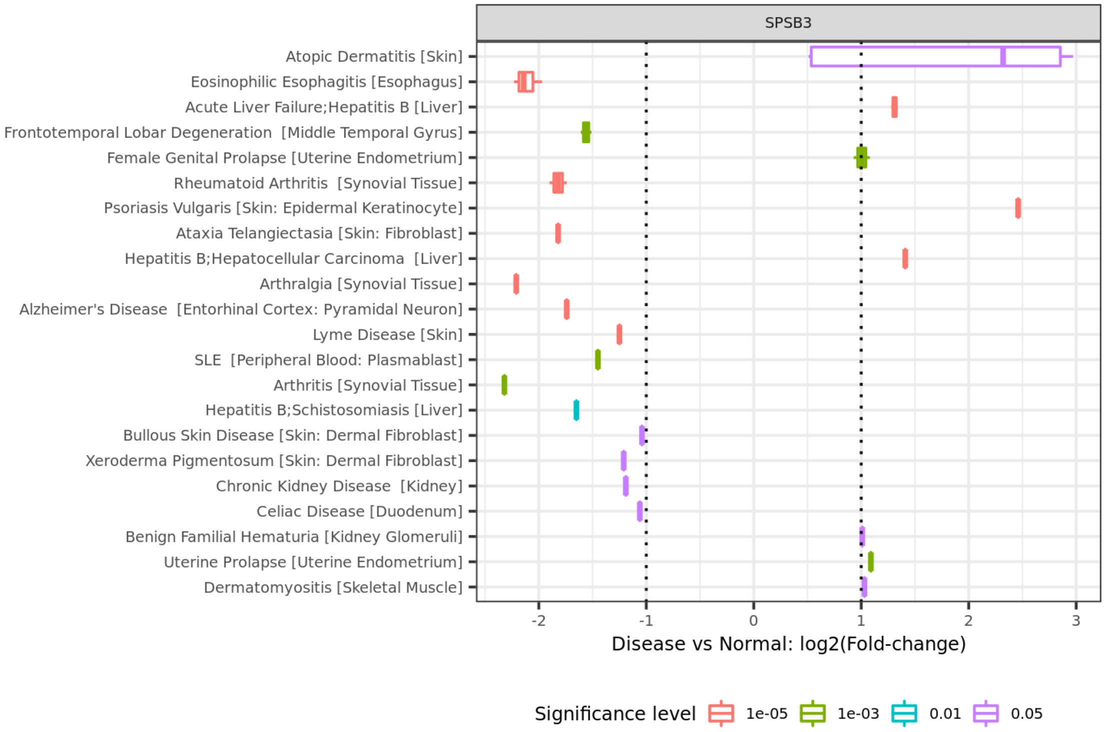
SPSB3
Atopic Dermatitis [Skin]
Eosinophilic Esophagitis [Esophagus]
Acute Liver Failure;Hepatitis B [Liver]
Frontotemporal Lobar Degeneration [Middle Temporal Gyrus]
Female Genital Prolapse [Uterine Endometrium]
Rheumatoid Arthritis [Synovial Tissue]
Psoriasis Vulgaris [Skin: Epidermal Keratinocyte]
Ataxia Telangiectasia [Skin: Fibroblast]
Hepatitis B;Hepatocellular Carcinoma [Liver]
Arthralgia [Synovial Tissue]
Alzheimer's Disease [Entorhinal Cortex: Pyramidal Neuron]
Lyme Disease [Skin]
SLE [Peripheral Blood: Plasmablast]
Arthritis [Synovial Tissue]
Hepatitis B;Schistosomiasis [Liver]
Bullous Skin Disease [Skin: Dermal Fibroblast]
Xeroderma Pigmentosum [Skin: Dermal Fibroblast]
Chronic Kidney Disease [Kidney]
Celiac Disease [Duodenum]
Benign Familial Hematuria [Kidney Glomeruli]
Uterine Prolapse [Uterine Endometrium]
Dermatomyositis [Skeletal Muscle]
-2
-1
0
1
2
3
Disease vs Normal: log2(Fold-change)
Significance level
1e-05
1e-03
0.01
0.05

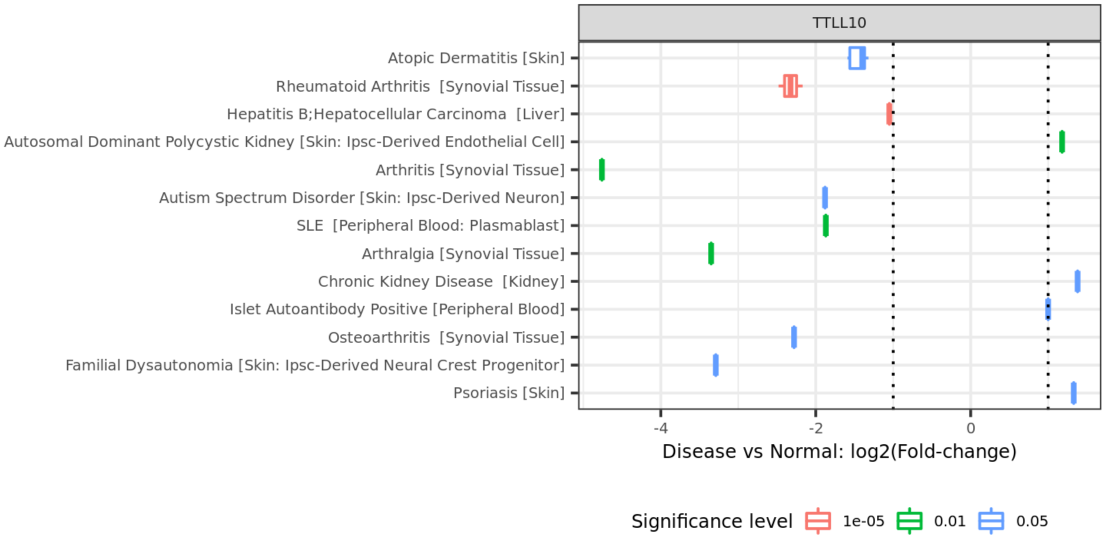


Supplementary figure 3. Differential expression results for severe acne related genes identified in MR and Colocalization analyses. Traits shown in this figure are significant ones passed multi-test adjustment (FDR < 0.05).
