## Supplementary figure 4 for "Genome-wide association study analysis of disease severity in Acne reveals novel biological insights"

### A) *ADAM23*

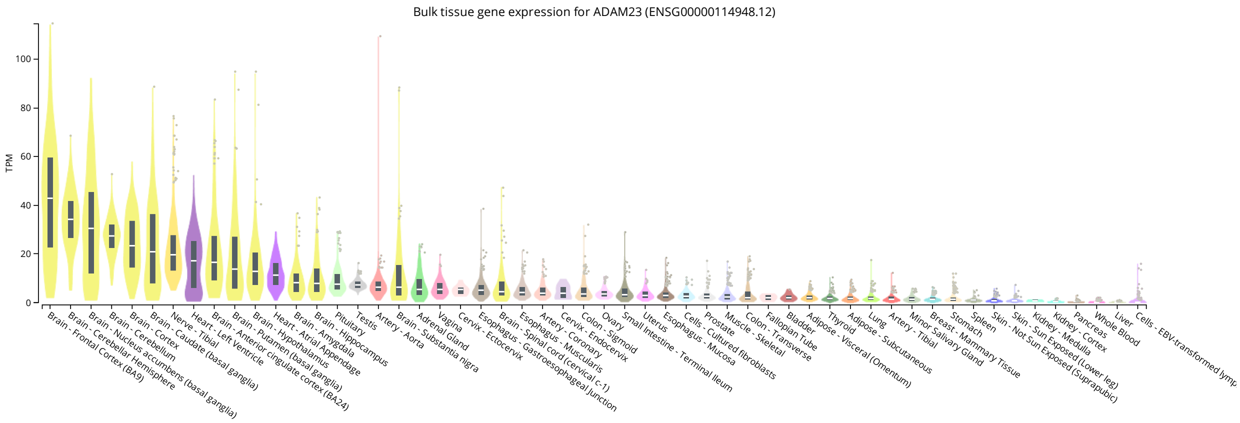


### B) *ATXN7L1*

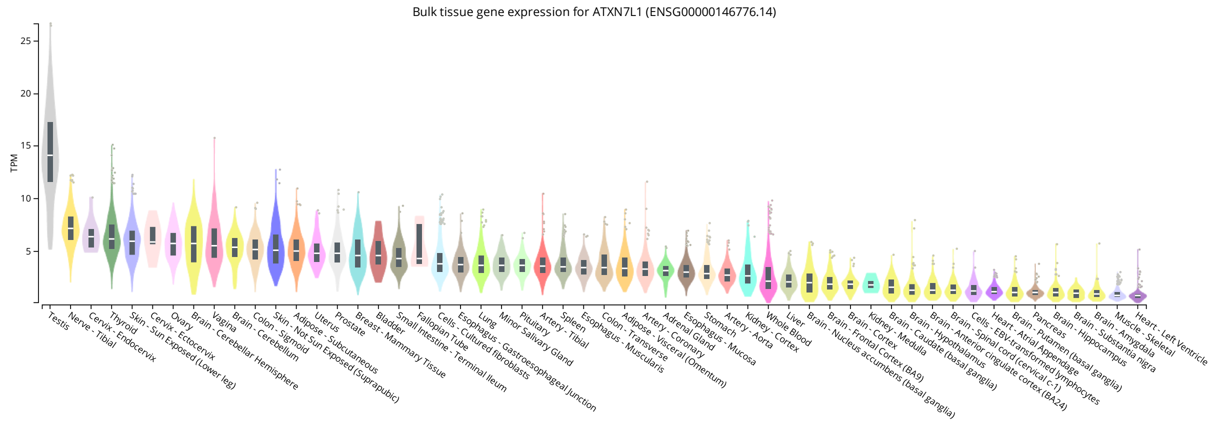


### C) *CDC7*

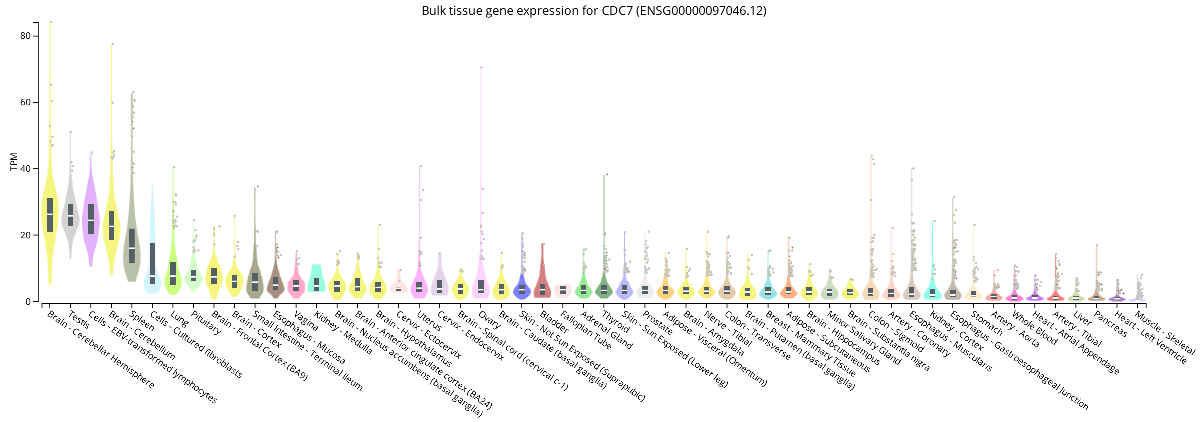


### D) *CDK20*

Confidential - Sensitive

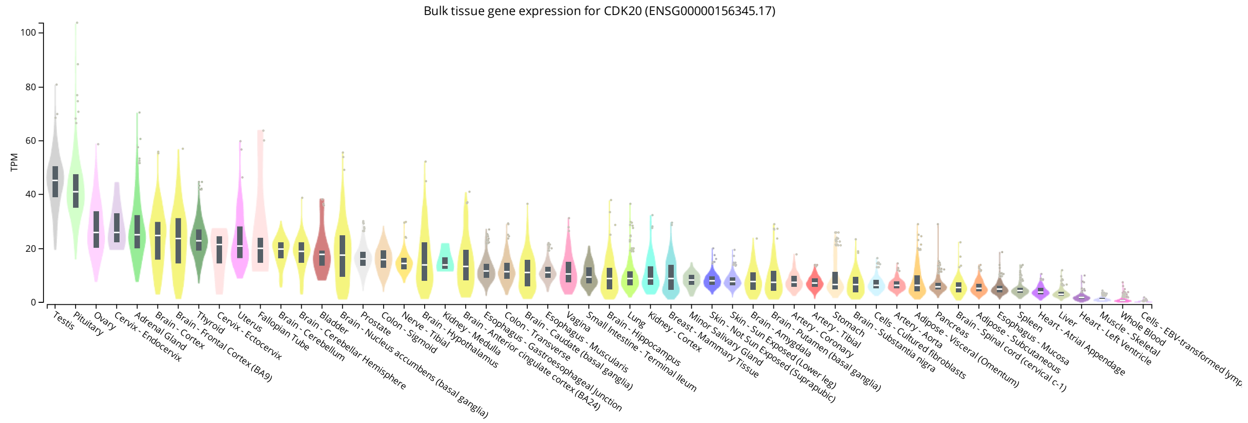


### E) *DNAJA4*

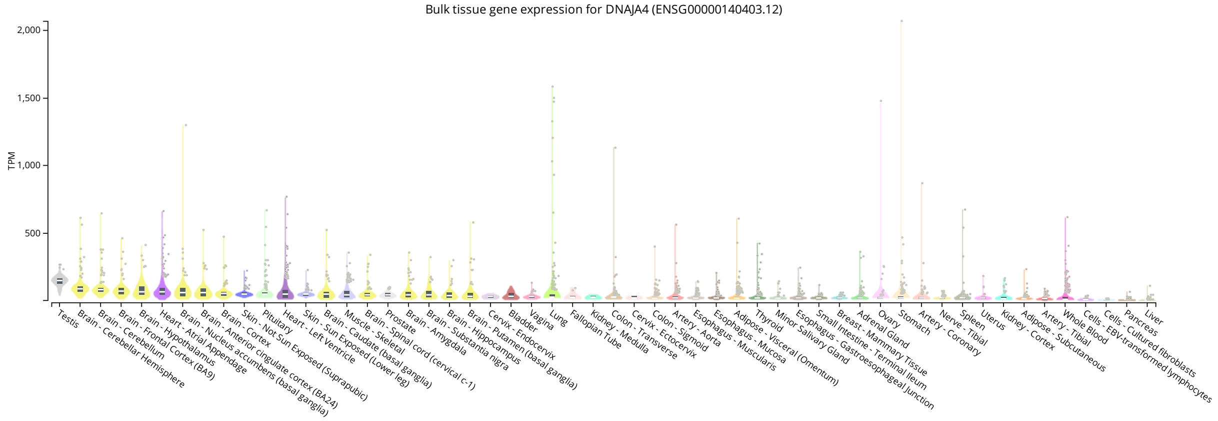


### F) *SLC7A1*

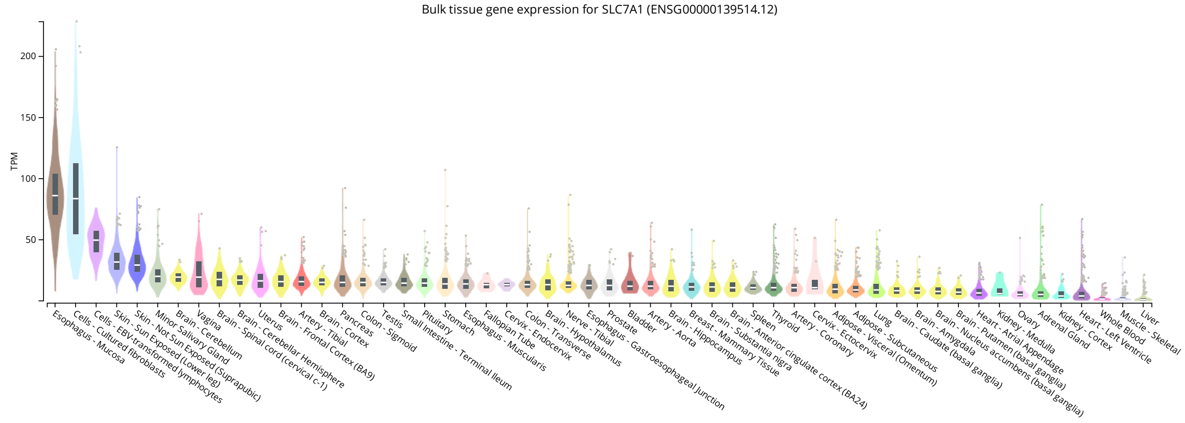


### G) *TTLL10*

Confidential - Sensitive

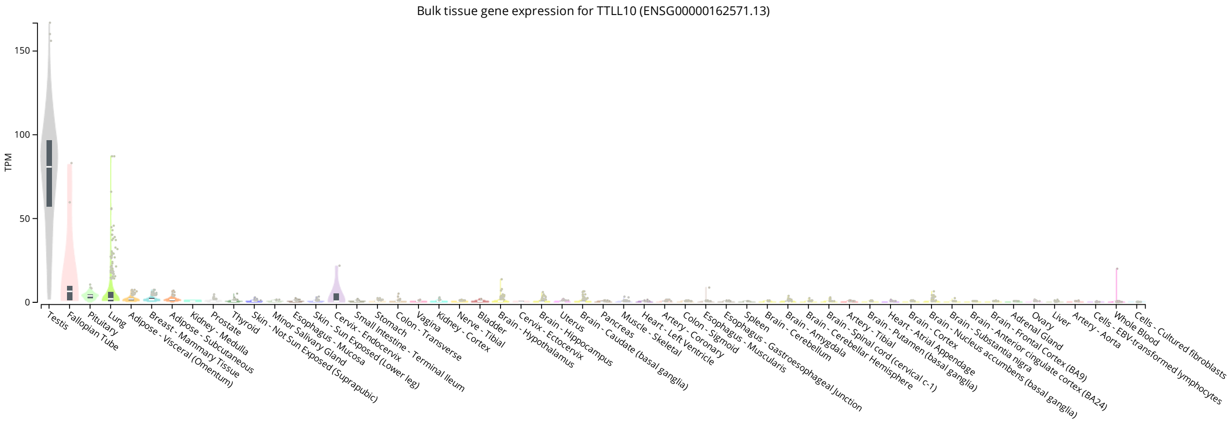


Supplementary figure 4. Bulk tissue-specific gene expression of severe acne related genes in GTEx database.
